# Improving the diagnosis of endometrial hyperplasia using computerized analysis and immunohistochemical biomarkers

**DOI:** 10.1101/2022.02.27.22271450

**Authors:** Peter A Sanderson, Arantza Esnal-Zufiaurre, Mark J Arends, C Simon Herrington, Frances Collins, Alistair RW Williams, Philippa TK Saunders

## Abstract

Endometrial hyperplasia (EH) is a precursor lesion to endometrial carcinoma (EC). Risks for EC include genetic, hormonal and metabolic factors most notably those associated with obesity: rates are rising and there is concern that cases in pre-menopausal women may remain undetected. Making an accurate distinction between benign and pre-malignant disease is both a challenge for the pathologist and important to the gynaecologist who wants to deliver the most appropriate care to meet the needs of the patient. Premalignant change may be recognised by histological changes of endometrial hyperplasia (which may occur with or without atypia) and endometrial intraepithelial neoplasia (EIN).

In this study we created a tissue resource of EH samples diagnosed between 2004 and 2009 (n=125) and used this to address key questions: 1. Are the EIN/WHO2014 diagnostic criteria able to consistently identify premalignant endometrium? 2. Can computer aided image analysis inform identification of EIN? 3. Can we improve diagnosis by incorporating analysis of protein expression using immunohistochemistry.

Our findings confirmed the inclusion of EIN in diagnostic criteria resulted in a better agreement between expert pathologists compared with the previous WHO94 criteria used for the original diagnosis of our sample set. A computer model based on assessment of stromal:epithelial ratio appeared most accurate in classification of areas of tissue without EIN. From an extensive panel of putative endometrial protein tissue biomarkers a score based on assessment of HAND2, PTEN and PAX2 was able to identify four clusters one of which appeared to be more likely to be benign.

In summary, our study has highlighted new opportunities to improve diagnosis of pre-malignant disease in endometrium and provide a platform for further research on this important topic.

**Highlights:**

1. Blinded re-analysis of n=125 samples previously classified as endometrial hyperplasia found improved intra-observer agreement (67%) using EIN/WHO2014 compared with standard WHO1994 criteria (52%)
2. Computerised analysis of endometrial hyperplasia tissue architecture showed promise as a tool to assist pathologists in diagnosis of difficult to classify cases
3. A diagnosis of endometrial intraepithelial neoplasia (EIN) using the WHO2014 criteria more accurately predicted risk of endometrial cancer than WHO1994 system.
4. EIN samples exhibited altered expression of ARID1A (negative glands) and HAND2 (reduced or absent from stroma)
5. Unsupervised hierarchical cluster analysis based on immunostaining for PTEN, PAX2 and HAND2 identified 4 subtypes one of which segregated with benign disease.
6. These results provide a framework for improved classification of pre-malignant lesions in endometrium that may inform personalized care pathways

## Introduction

Endometrial hyperplasia (EH) is an ‘umbrella’ term that incorporates a heterogeneous spectrum of abnormal endometrial lesions (Sanderson et al., 2017). The clinical significance of a diagnosis of endometrial hyperplasia lies in its association with an increased risk of progression to the endometrioid subtype of endometrial carcinoma (EC). Endometrial carcinoma is the most common gynaecological malignancy in the UK with ∼9K cases per year [https://www.cancerresearchuk.org/about-cancer/womb-cancer/about]. Type I endometrioid EC’s account for ∼75% of cases with unopposed oestrogen action implicated in their aetiology (Sanderson et al., 2017). A revised subtype analysis of endometrial cancers based on genetic changes (Kandoth et al., 2013) has highlighted the importance of broadening the criteria for evaluation of EH samples to refine the association with risk of progression to malignancy.

Diagnosis of EH or EC in postmenopausal women is most often triggered by an experience of uterine bleeding and historically EHs were estimated to account for 15% of all cases of postmenopausal bleeding (Lidor et al., 1986). Two of the high-risk patient populations prone to the development of EH are (i) obese peri/postmenopausal women, due to peripheral aromatisation of androgens to oestrogens in adipose tissue, coupled with erratic anovulatory menstrual cycles and (ii) premenopausal patients with polycystic ovarian syndrome (PCOS), due to hyperandrogenic anovulation. National Guidelines published in the UK [https://www.rcog.org.uk/en/guidelines-research-services/guidelines/gtg67/] and other countries [https://www.jogc.com/article/S1701-2163(19)30452-9/fulltext] emphasise the need to customise them taking into account baseline risk factors, symptomatology, fertility wishes and response to treatment). Three management options are usually considered: active surveillance, progestin therapy or hysterectomy. Treatment in pre-menopausal women is usually focused on medical management rather than surgery (Pennant et al., 2017). EHs occurring entirely due to unopposed oestrogen exposure, i.e. an ‘endocrine effect’, may be capable of regression back to normal endometrium through the withdrawal of the oestrogen source or using exogenous progesterone administration to oppose the impact of oestrogens and reduce epithelial cell proliferation. Progestin therapy has been demonstrated to be effective by multiple studies in achieving regression of endometrial hyperplasia (Gallos et al., 2013). Regional practice varies on the route of progestin administration, however both continuous oral and local intrauterine (levonorgestrel-releasing intrauterine system (LNG-IUS) are reported to be effective in achieving regression although there is a higher incidence of failure when cytological atypia is present (Gallos et al., 2012).

Based on reports that metformin can also reduce endometrial cell proliferation this drug has been explored as an alternative to progestins as they can cause breakthrough bleeding (Clement et al., 2017). In a recent systematic review Chae-Kim et al reported that reproductive-aged women with atypical EH or early endometrial cancer, had lower relapse rates when treated with combined progestin and metformin compared with progestin therapy alone, with similar pregnancy rates highlighting the potential for fertility sparing treatments in this age group (Chae-Kim et al., 2021).

High rates of abnormal pathology have consistently been reported in endometrial biopsies from morbidly obese women (Kaiyrlykyzy et al., 2015). Several studies have reported the normalisation of endometrial pathology in obese women following bariatric surgery, improved response to progestin therapy and reduced cancer risk (MacKintosh et al., 2019, Barr et al., 2021, Argenta et al., 2014). Weight loss is not always associated with complete normalisation of pathology and it has been suggested monitoring should be continue and histological screening might be justified.

A challenge for gynaecologist and pathologist alike is the reproducible stratification of women with EH attributable to purely endocrine factors, for example as a result of chronic unopposed oestrogen exposure, from those women with EH where the tissue has malignant potential. One of the most obvious characteristics of EH tissue is the presence of excess/irregular proliferation within the glandular epithelial compartment which can be seen as a change in the endometrial gland-to-stroma ratio compared to endometrium from the normal proliferative phase of the menstrual cycle. Whilst the appearance of the glands may also vary the presence of cells with abnormal shape/size and nuclear architecture (cytological atypia) is generally accepted as a histological characteristic that predicts progression to malignancy (Kurman et al., 1985).

Whilst several different classification systems have been developed there are three that have been used most extensively. In 1994 the World Health organisation recommended a four-tier classification system that considered cytological and architectural abnormalities within EH lesions, categorising them into 4 types: simple without nuclear atypia (SH), simple with atypia (SAH), complex without atypia (CH) and complex atypical hyperplasia (CAH). Despite extensive use and popularity within modern gynaecological practice, the WHO1994 system has been challenged as it can fail to deal with the heterogeneity demonstrated by EH lesions and to align with the therapeutic options available (Baak and Mutter, 2005). An alternative classification was proposed based around the molecular assessment of the clonality of EH lesions and a shared lineage between premalignant EH lesions and the cancers that develop from them (Mutter et al., 1996, Mutter, 2002). This endometrial intraepithelial neoplasia (EIN) classification system divides potential lesions into two groups: (i) benign EH and (ii) EIN (the premalignant lesion). EIN classification categories do not correspond directly to specific categories in the WHO94 system although there is an element of recognisable overlap. In 2014 the WHO published a new edition of their classification (hereafter referred to as EIN/WHO2014) which changed their recommendations on classification so that they were more obviously aligned with those proposed for EIN and reduced the EH classification into a) hyperplasia without atypia (HwA) b) atypical hyperplasia/EIN (Kurman et al., 2014).

The primary objective of the current study was to use histological evaluation in combination with computer aided analysis and immunostaining for more reliable diagnosis of premalignant EH. To achieve this objective we established a tissue resource retrieved from the pathology department of NHS Lothian which consisted of 125 endometrial samples which were originally diagnosed as EH in 2004-2009 based on the WHO94 classification system. We selected targets for immunohistochemical analysis based on proteins encoded by genes implicated in progression to endometrial malignancy (p53, PTEN, PAX2, ARID1A) (Baak et al., 2005b, Jarboe et al., 2009, Monte et al., 2010), mismatch repair processes (MLH1, MSH1, MSH6, PMS2) (Esteller et al., 1999, Crosbie et al., 2018) and stromal-epithelial regulation of endometrium (HAND2) (Li et al., 2011). We confirmed application of the WHO 2014/EIN criteria resulted in greater diagnostic concordance between pathologists and that computer aided evaluation of digitised images was also beneficial in diagnosis of EIN. We detected altered patterns of expression of ARID1A and HAND2 that correlated with presence of EIN and were able to cluster samples based on immunostaining for PTEN/PAX2/HAND2 as a first step towards improving information that might inform personalised care.

## Materials and Methods

### Establishment of an endometrial hyperplasia tissue resource

No primary tissue samples were collected during this study. Analysis was undertaken using samples of endometrium archived within the Pathology Department of the NHS Lothian Health Board that had been recovered during routine surgery. These samples are managed within the Lothian NRS Human Annotated Bioresource that was granted ethical approval by the East of Scotland Research Ethics Service (REC 1) in 2015 [https://www.hra.nhs.uk/planning-and-improving-research/application-summaries/research-summaries/lothian-nrs-human-annotated-bioresource/]. Copies of the sample application and approval are shown in Supplementary Figures 1 and 2.

The EH samples for the study were identified as follows: following a search of the NHS Lothian pathology ‘Apex’ clinical database (by PAS and ARWW), n=143 patient cases clinically coded with a diagnosis of EH between January 2004 and December 2009 were identified. After exclusions and accounting for the availability of archival tissue, n=125 EH patients were identified (Figure 1). Serial sections of formalin-fixed, paraffin embedded tissue (n=125 identified as above) were obtained together with anonymised information on patient demographics and medical history.

**Figure 1.**
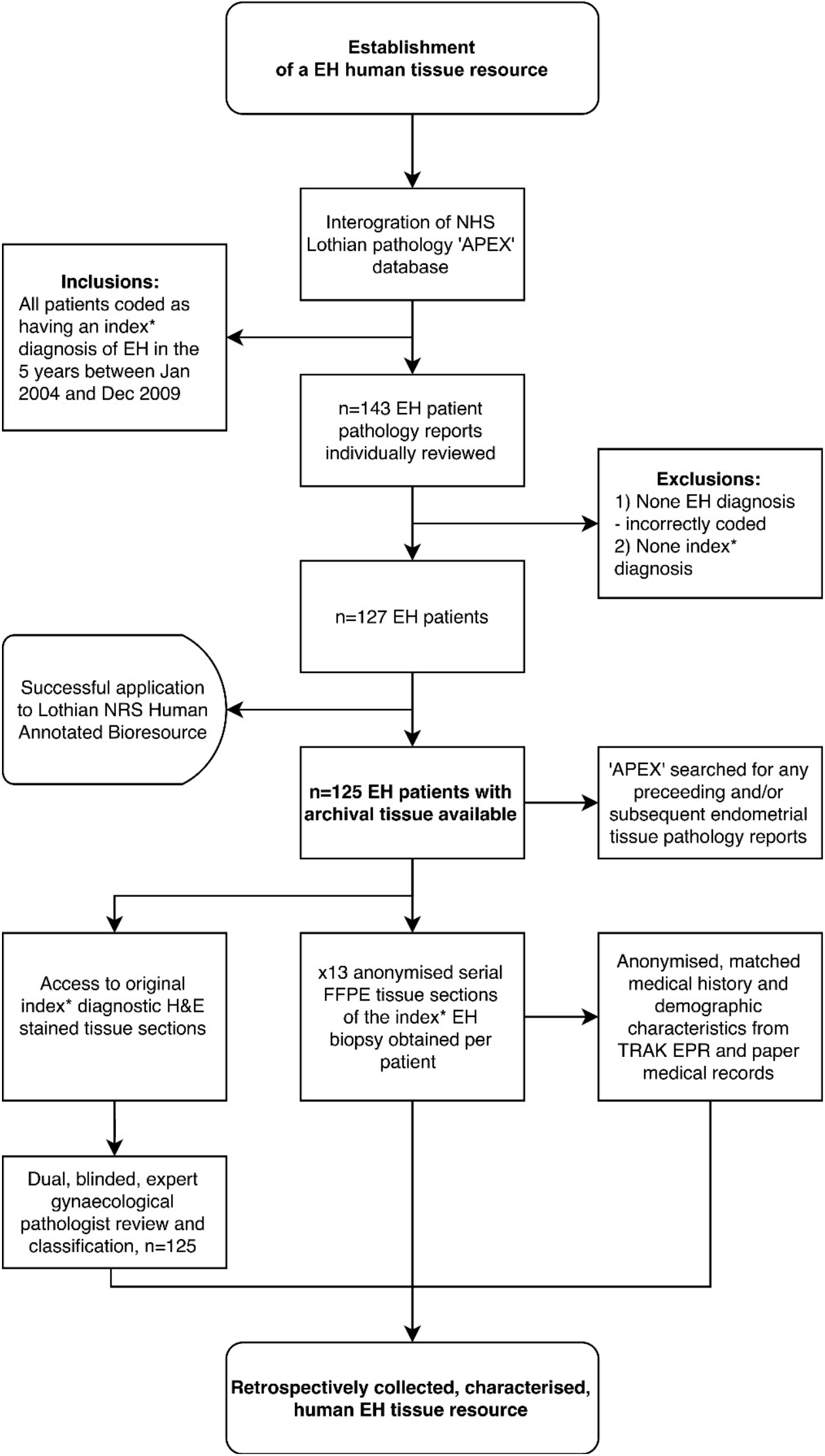
Summary: Workflow-diagram for the establishment of a human endometrial hyperplasia (EH) tissue resource. *Index refers to the first documented EH biopsy, i.e. not a repeat or follow-up biopsy. TRAK EPR = NHS Lothian’s electronic patient records system. APEX = NHS Lothian’s pathology records system

### Histological assessment, reclassification and imaging

EH tissue samples underwent a dual, blinded, re-review by two expert gynaecological pathologists (ARW and SH) using utilising a standardised diagnostic proforma (Supplementary Figure 3). Each pathologist undertook an evaluation of the sample set according to both the WHO1994 and EIN/WHO2014 classification systems. No clinical information was provided at the time of the review to blind the pathologists and reduce bias.

Where there was a diagnostic discrepancy between the two expert pathologists using the EIN/WHO2014 system, both were asked to re-review the discrepant samples using a dual-headed microscope to reach a final consensus diagnosis. A third independent pathologist was asked to settle any unresolvable discrepancies. For the purposes of the consensus review, only those discrepancies that would hypothetically result in a change to clinical management (n=32) were subject to re-review, e.g., where one pathologist diagnosed a case as HwA and the other pathologist diagnosed the same case as EIN.

Where there were discrepancies between assignment as disordered proliferative endometrium or HwA, cases were upgraded into the HwA category. Discrepancies between two benign diagnoses were upgraded to the more ‘abnormal’ of the two in order to form the final diagnosis, e.g. a discrepancy between proliferative endometrium and disordered proliferative endometrium was assigned as the latter.

### Digital computerised quantitative image analysis

As a complement to the pathologist re-review all H&E slides were also scanned using NanoZoomer-XR scanner in 40x mode and stored as NanoZoomer Digital pathology files (.pdpi). Scanned H&E sections were examined by ARW and PAS and on each slide two regions of interest (ROI) were identified and digitally marked. One ROI corresponded to the pathologically ‘most abnormal’ appearing area of the sample. For example, in samples with a consensus expert pathologist diagnosis of EIN, the ‘most abnormal’ ROI corresponded to the entire clonal expansion of EIN (or where the entire tissue section contained only EIN / multiple foci, then the most abnormal focus of EIN was marked). The second ROI marked within the sample corresponded to the background endometrium or the ‘least abnormal’ area. Where the sample had a non-EIN consensus diagnosis, the ‘most abnormal’ ROI corresponded to an area displaying the most representative pathological features of the non-EIN areas, whilst the ‘least abnormal’ ROI within the sample corresponded to the background endometrium.

The ROI were evaluated using the StrataQuest analysis software, v5.0 (TissueGnostics GmbH, Vienna). Pre-defined, bespoke analysis parameters for endometrial H&E image processing and pattern recognition algorithms were commercially designed in ‘the H&E app’ (TissueGnostics GmbH, Vienna) and applied to individual regions of interest (ROI) within the imported EH images. After manual optimisation and correction of analysis parameters, layered segmentation or ‘masks’ were applied. These were assigned colours (see example in Supplementary Figure. 4) by the software dependent on the tissue structure being detected (i.e. endometrial stromal cells = dark green). On final analysis the ‘masks’ were built up to give a final image and numerical data calculated for each tissue compartment, e.g. tissue area, number of nuclei, etc. Volume percentage stroma (VPS) was calculated as: VPS = total stromal area (dark green) / [total stromal area (dark green) + total epithelial area (blue + red) + total glandular lumen area (light green) + total vessel area (purple)] x 100.

### Immunohistochemistry

Immunohistochemistry was performed using standard protocols established within the laboratory using commercially available ImmPRESS detection kits (Vector Laboratories, Inc., Burlingame, USA). The kits contain a ready-to-use ImmPRESS reagent, which employs horseradish peroxidase (HRP) micropolymers conjugated to affinity-purified secondary antibodies. This permits a higher density of enzymes per antibody to bind to the target, increases binding specificity and reduces background staining. Positive bound antibodies were detected using 3, 3 – diaminobenzidine (DAB), counterstained with haemotoxylin and mounted in Pertex (Cellpath Technologies, UK). Details of primary antibodies, their supplier, catalogue number and dilution are in Supplementary Table 1.

### Scoring of immunohistochemistry

Evaluation of immunohistochemical staining patterns was carried out by a minimum of 2 of the co-authors unless specified using the entire section of tissue in all cases.

#### P53

Staining was considered ‘wild-type’ if it was patchy/heterogeneous but aberrant if absent from nuclei or intense in cytoplasm (nuclear staining present or absent) (Kobel et al., 2016).

#### PTEN

Staining was scored using a modified method based on Mutter et al (Mutter et al., 2000). Scores of 0, 1 and 2 were allocated based on the presence (0, glands immunopositive) or absence (1, isolated immunonegative (null) glands; 2 focal area, >2 null glands) cytoplasmic/nuclear staining in epithelial cells (examples of staining patterns are illustrated in Figures 6 and 7 of (Sanderson et al., 2017).

#### PAX2

Staining was scored according to criteria of Quick et al (Quick et al., 2012). Positive (score 0) all glands with nuclear staining, ‘null’ (score 1) if small, isolated, groups of glands had no staining and ‘altered (score 2) if large areas or all glands were immunonegative.

#### Mismatch repair proteins

Immunohistochemistry for the DNA Mismatch repair proteins (MLH1, MSH2, MSH6 and PMS2) were scored in EH tissues as described by Woo et al (Woo et al., 2014) and in keeping with UK NEQAS recommendations (Arends et al. 2008; [https://www.acgs.uk.com/media/10772/hnpcc_recommendations_b.pdf] Normal human vermiform appendix tissue was used as a positive control.

#### HAND2

Scoring for HAND2 staining was undertaken by 3 independent members of the Saunders’ laboratory (IS, PK, OK) not otherwise involved in the processing or evaluation of samples who were asked to score staining in the two ROI of each section based solely on the amount of staining in the stromal compartment. A score of 0 (absent expression) was given if 0 % of stromal nuclei in the designated area stained brown, 1 (reduced expression) if 1-50 % of the stromal nuclei in the designated area stained brown and 2 (positive) if >50% of stromal nuclei in the designated area stained brown. The scoring results of ‘PK’ and ‘OK’ were compared for consensus agreement. Where a consensus was not reached, the score from ‘IS’ was used to achieve a 2/3 majority consensus.

#### ARID1A

Scoring used a modified method based on (Ayhan et al., 2015): a score of ‘positive’ was assigned if both glands and stroma were immunopositive. Stromal staining was detected in all samples (serving as an internal control) whereas glands were sometimes immunonegative ranging from focal loss (groups of adjacent negative glands) to complete loss of expression in all glands even if the adjacent stroma was positive.

### Statistical analysis

Statistical analysis was performed using GraphPad Prism 8.0. The two-sided Fisher’s Exact test was used to compare between groups for the immunostaining data.

### Unsupervised hierarchical agglomerate clustering

Unsupervised hierarchical agglomerative clustering was used to evaluate the correlation of immunohistochemical scoring data with EH diagnosis and any recorded malignant progression. Clustering analysis organises the data according to the similarity/dissimilarity of immunostaining profiles, arranging the cases with similar immunoprofiles together in rows in a heatmap. The relationship between EH cases and immunomarkers is displayed graphically as a dendrogram, where the branch length is determined by correlation between immunostaining scores.

Immunohistochemical score data was formatted as described by Liu and colleagues (Liu et al., 2002) followed by analysis and visualisation using the Cluster and TreeView software platforms respectively, as described by Eisen, et al (Eisen et al., 1998). The clustering of the immunohistochemical data was performed using the complete linkage method and the Euclidean distance. Comparison was performed to the average linkage clustering method to assess reproducibility of the cluster groups described. This demonstrated a 75.2 % agreement with a Kappa, *k* score of 0.629 “Substantial”. Chi-squared and Fisher’s exact tests were used to determine which EH diagnosis and immunohistochemical markers contributed to the formation of individual clusters.

## Results

### Interobserver variability was apparent when diagnosing EH using the WHO94 classification

All 125 EH lesions were originally diagnosed and coded by NHS pathologists between 2004 and 2009 utilising the WHO94 classification system. (Table 1 index diagnosis). Results from a blinded re-review by two expert pathologists (A and B) using the same criteria is also displayed in Table. 1. For all 125 cases, the percentage agreement between pathologists A and B and the original index diagnosis was 56.0 % (n=70) and 48.8 % (n=61) respectively. This amounted to a Cohen’s Kappa (*k*) interobserver agreement score of ‘fair’ for pathologist A and ‘slight’ for pathologist B. Of note the complex hyperplasia (CH) category exhibited the lowest levels of agreement between the index diagnoses and the re-review with pathologist A agreeing with 12/29 (41.3 %) of the index diagnoses and pathologist B not agreeing with any. Pathologist B upgraded two diagnoses from EH to malignancy.

**Table 1.**
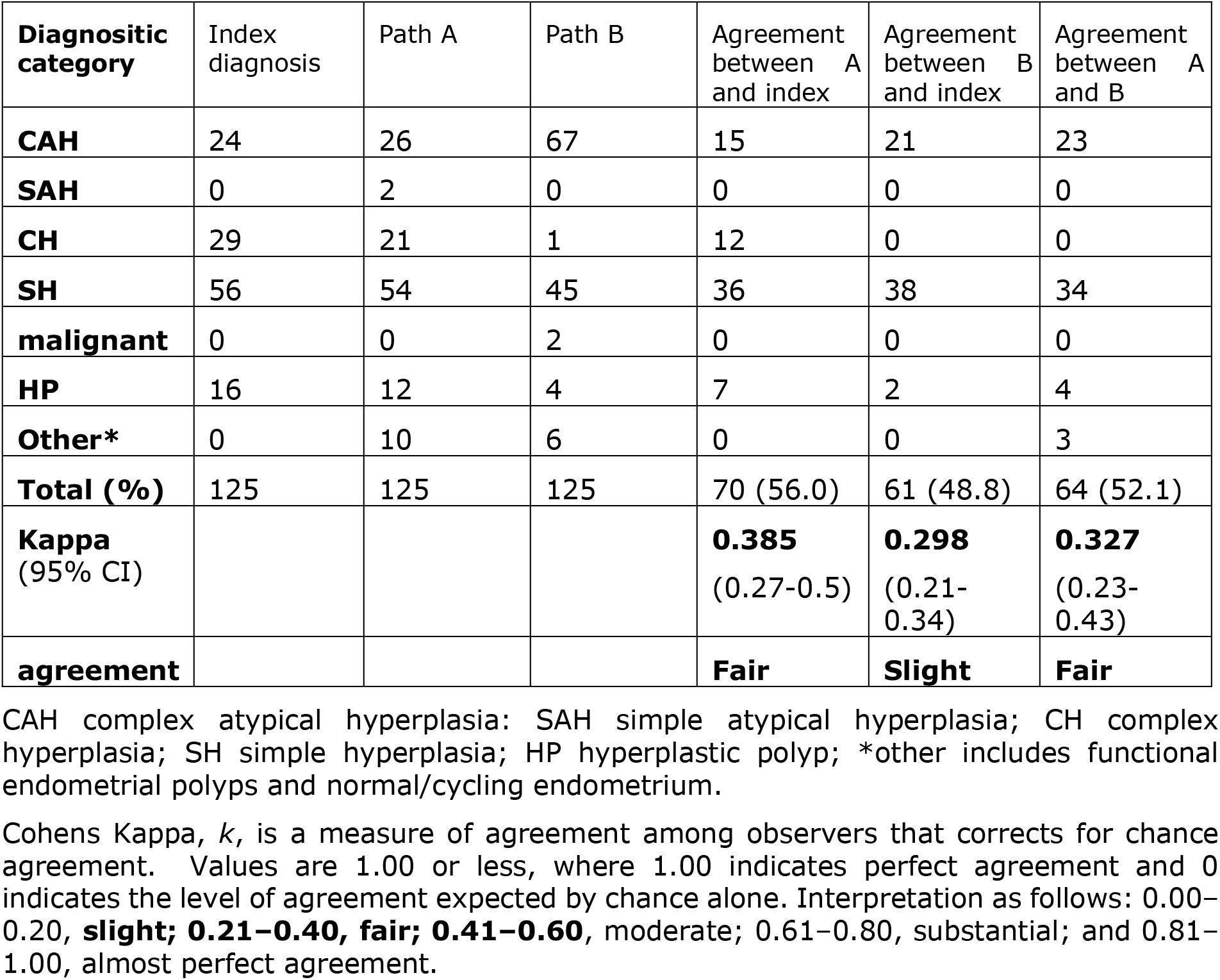
Comparison between endometrial hyperplasia diagnosis by independent pathologists using WHO94 classification revealed discordant findings. Numbers in each column show results of evaluation using a H&E-stained section of endometrial tissue from n=125 women.

Interobserver agreement was assessed but agreement was no better than when comparison was made with the original index diagnoses, with total percentage agreement reaching 52.1 % (n=64), *k* = 0.327 (95 % CI 0.23-0.43) ‘fair’.

Of particular concern was the inconsistency in diagnosis of the CAH which may have been compounded by the subdivision of this category in the original index diagnosis into six descriptive variants (Supplementary Table 2). As diagnosis of CAH can lead to a clinical recommendation of a hysterectomy there is concern that inconsistencies in the diagnosis of this patient subset could result a significant change in practice.

### Evaluation of samples in the EH tissue resource using the EIN/WHO2014 system results in a more consistent diagnosis and improved intra-observer agreement

Using the updated 2014 criteria the agreement between the diagnoses from expert pathologist was higher than that previously seen when utilising the WHO94 system, standing at 67.2 % (n=84) and amounting to an interobserver agreement score of *k* = 0.478 (95 % CI 0.356-0.600) ‘moderate’ (Table 2). Interestingly, and somewhat unexpectedly, pathologist A diagnosed n=46 cases of EIN and pathologist B diagnosed n=66 cases, both noticeably higher than the number of cases originally given an index diagnosis of CAH (n=24). Comparison between the index cases of CAH and those reclassified with EIN revealed an overlap of n=20 samples with 3 reclassified as HwA and one as malignant. Whilst 1^st^ line treatment of hysterectomy would be appropriate for 21 of these patients 3 may have been offered surgery when medical management might have been appropriate. Of greater concern is the 32 patients diagnosed with lesions considered less likely to progress to malignancy (CH/SH) who might have benefitted from surgical treatment.

**Table 2.**
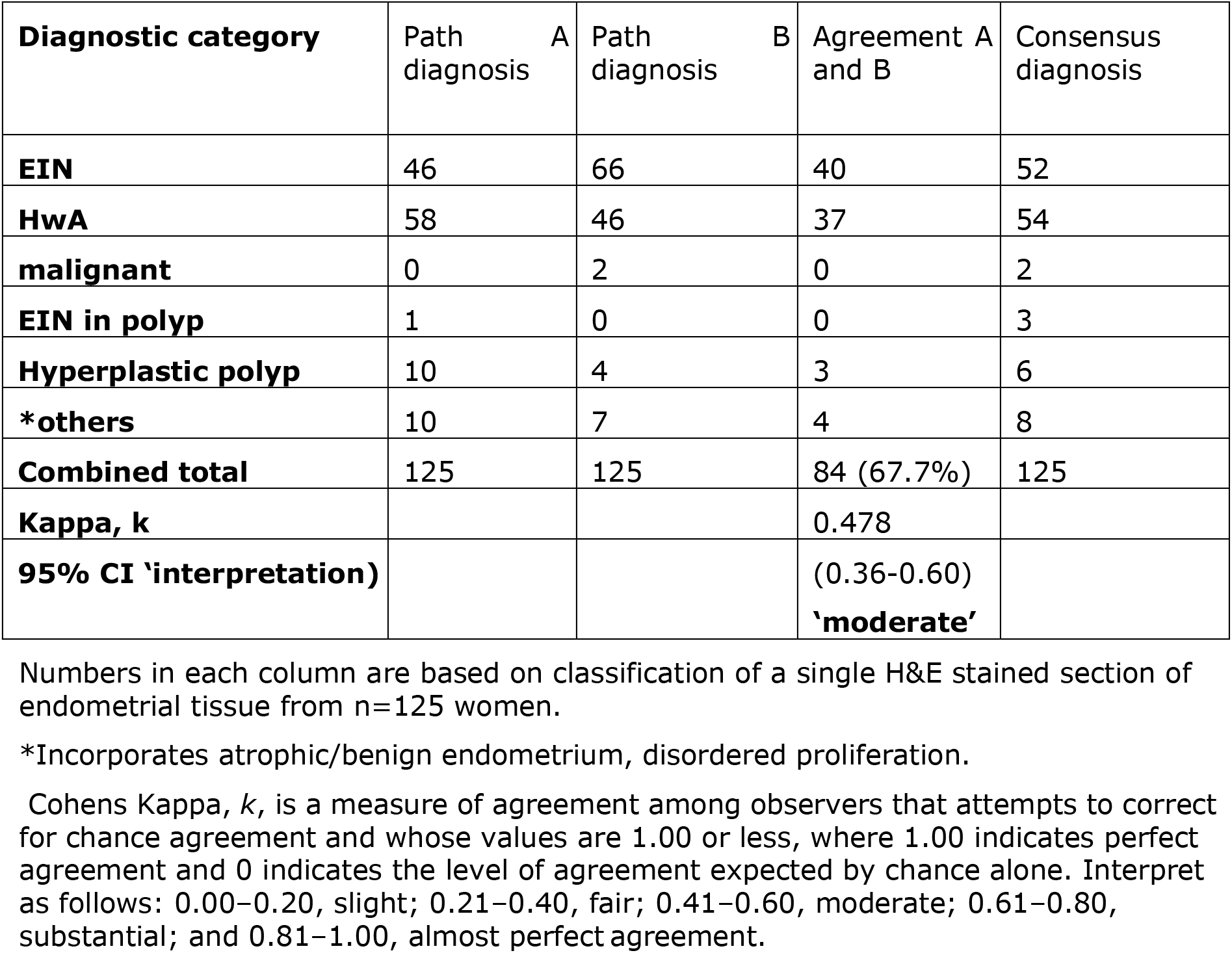
Interobserver agreement with diagnosing endometrial hyperplasia using the EIN/WHO 2014 classification.

| **Diagnostic category** | Path diagnosis A | Path diagnosis B | Agreement A and B | Consensus diagnosis |
| --- | --- | --- | --- | --- |
| **EIN** | 46 | 66 | 40 | 52 |
| **HwA** | 58 | 46 | 37 | 54 |
| **malignant** | 0 | 2 | 0 | 2 |
| **EIN in polyp** | 1 | 0 | 0 | 3 |
| **Hyperplastic polyp** | 10 | 4 | 3 | 6 |
| **\*others** | 10 | 7 | 4 | 8 |
| **Combined total** | 125 | 125 | 84 (67.7%) | 125 |
| **Kappa, k** |  |  | 0.478 |  |
| **95% CI 'interpretation)** |  |  | (0.36-0.60)<br>**'moderate'** |  |

Numbers in each column are based on classification of a single H&E stained section of endometrial tissue from n=125 women.
\*Incorporates atrophic/benign endometrium, disordered proliferation.
Cohens Kappa, *k*, is a measure of agreement among observers that attempts to correct for chance agreement and whose values are 1.00 or less, where 1.00 indicates perfect agreement and 0 indicates the level of agreement expected by chance alone. Interpret as follows: 0.00–0.20, slight; 0.21–0.40, fair; 0.41–0.60, moderate; 0.61–0.80, substantial; and 0.81–1.00, almost perfect agreement.

A summary of the demographics of the two groups of patients reclassified as EIN (n=52) or hyperplasia without atypia (HwA, n=54) is given in Supplementary Table 3. There were no significant differences in age, ethnicity or menopausal status; patients with EIN were less likely to have had children and to have a diagnosis of PCOS.

### The EIN/WHO classification system more often correctly predicted a malignant outcome

Twelve (10.17 %) from 118 (n=7 lost to follow-up from the original n=125 cohort) patients for whom the index endometrial biopsy was classified as EH were later diagnosed with an endometrioid EC. Median time from index EH diagnosis to EC diagnosis was 146.5 days (range 36 – 3481 days, standard deviation, SD 1081.46 days). Ten of the ECs were diagnosed within 12 months of the index EH diagnosis and were therefore thought to represent concurrent cancers that were not sampled by the initial index endometrial biopsy. The remaining 2 ECs were diagnosed 1571 and 3481 days respectively after the initial index EH diagnosis and therefore developed subsequently. Of note, 5 of the patients who developed EC were premenopausal (41.7 %) and two were under 40 years of age.

Whilst the dataset was not large as a follow up to these analyses Kaplan-Meier ‘survival-curves’ detailing the percentage (y-axis) of patients with EH remaining cancer-free during the follow-up period (x-axis), are shown in Figure 2. The median (mean, SD) follow-up period was 3485 days (3180 days, 1383 days). Statistical analysis of the curves using a Log-rank (Mantel-Cox) test demonstrated a statistically significant difference between the curves for EIN vs. non-EIN (10.41, p=0.0013**), compared with CAH vs. non-CAH (2.115, p=0.145 ns). These results demonstrated that in this study an EIN diagnosis, as part of the EIN/WHO2014 classification system, better identifies those at risk of future EC than the CAH category in the WHO94 classification system: an EIN diagnosis carried a 13x higher chance of a subsequent or concurrent EC than a non-EIN diagnosis (Hazard ratio (HR) 13.37, 95 % CI 4.05-44.13) over the follow-up period, compared to a CAH diagnosis which had a 3x higher chance of a subsequent or concurrent EC than a non-CAH diagnosis (Hazard ratio, (HR) 3.029, 95 % CI 0.68-13.49).

**Figure 2.**
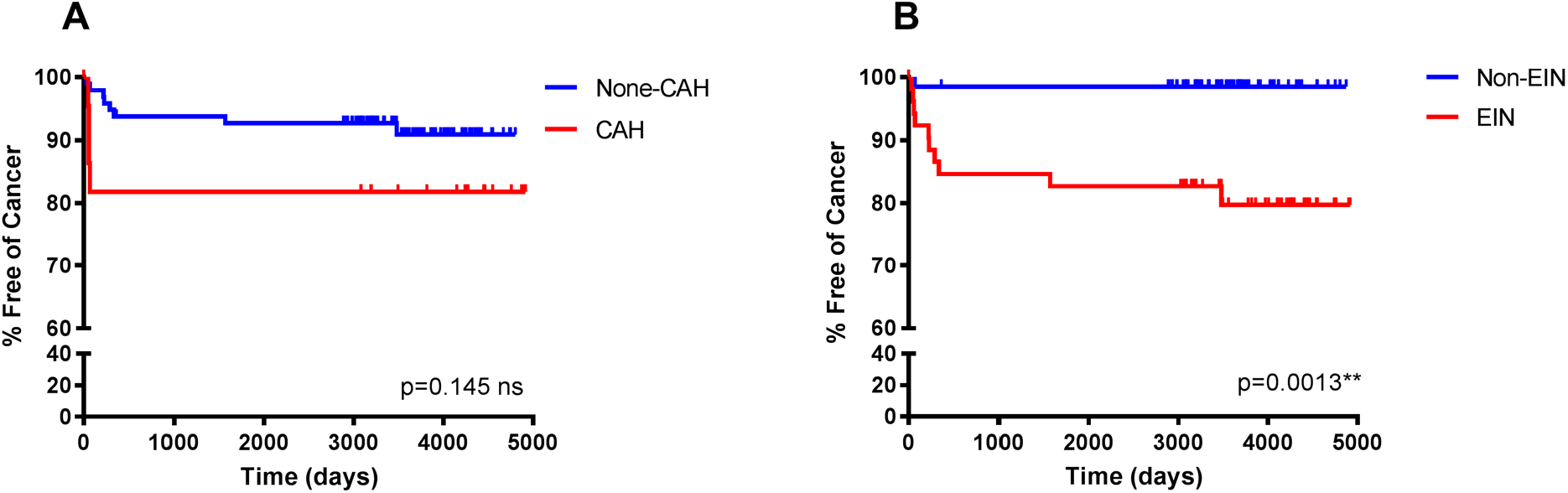
Kaplan-Meier curves to demonstrate the percentage of endometrial hyperplasia patients free of cancer during follow-up when classified using the WHO94 compared with the EIN/WHO2014 classification systems. Median follow-up period (mean, SD) 3485 days (3180 days, 1383 days). A) The original diagnostic classification of the patients in this study utilised WHO94 criteria. Percentage cancer-free time was not statistically significantly different between those with a WHO94 diagnosis of CAH vs those with a non-CAH diagnosis, Log-Rank (Mantel-Cox) 2.115, p=0.145 ns, HR 3.029 (95% CI 0.68-13.49). B) Consensus reclassification of the same EH cohort of patients utilising EIN/WHO2014, revealed a statistically significant difference in the percentage cancer-free time between those reclassified to an EIN diagnosis vs those reclassified to a non-EIN diagnosis, Log-Rank (Mantel-Cox) 10.41, p=0.0013**, HR 13.37 (95% CI 4.05-44.13). NB/ Non-CAH incorporates patients with CH (n=29), SH (n=51) and HP (n=15). EIN incorporates EIN (n=49) and EIN-EMP (n=3). Non-EIN incorporates patients with HwA (n=50) HP without EIN (n=6) and atrophic/benign endometrium (n=8).

### Computerised analysis of tissue compartments may assist pathologists with difficult to diagnose EH cases

To evaluate the use of semi-automated computer image analysis as a diagnostic adjunct to pathological classification, consensus EH cases (n=21) underwent digital image analysis to quantify the volume percentages of the stromal and epithelial tissue compartments. The ROIs deemed the ‘most abnormal’ within each tissue sample were used for the analysis. For the EIN samples (n=10) the ‘most abnormal’ ROI corresponded to a clonal region of EIN and for the HwA samples (n=11), the ‘most abnormal’ ROI corresponded to the most representative region of HwA.

Computerised digital quantification of the stromal and glandular compartments demonstrated that the consensus EIN cases (n=10), which by definition should have a VPS of <55 %, were identified by computer-assisted image analysis as having a VPS of <55 % in 30% (3/10) of cases suggesting that in the ‘most abnormal’ region of the tissue sections, 7/10 of the cases did not have glandular area which exceeded that of the stromal area by image analysis. Based on this image analysis evaluation of architecture alone these cases may not be considered as meeting the criterion for EIN as per the classification system (Baak and Mutter, 2005, Mutter, 2000). All the consensus cases of HwA (n=11) met the architectural requirements of the EIN/WHO2014 classification system and demonstrated a VPS of >55 % using this image analysis technique confirming that it could be used to validate exclusion of EIN (Figure 3).

**Figure 3.**
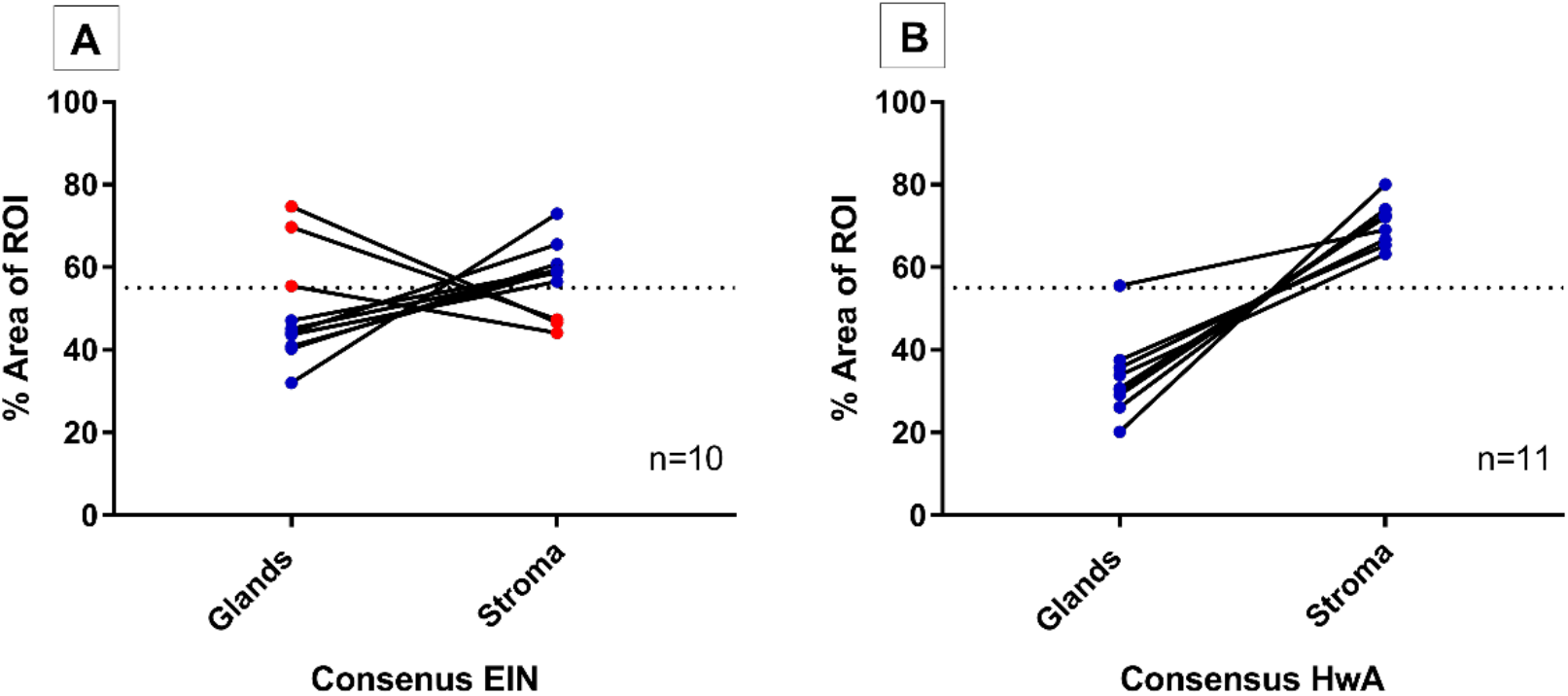
Semi-automated quantitative image analysis reveals differences between the final consensus pathology diagnosis and the EIN diagnostic criteria. Dashed line represents the 55 % threshold for volume percentage stroma (VPS) that is considered diagnositic. A) 3/10 (red) cases with a final consensus diagnosis of EIN met the EIN diagnostic criteria for architecture with a VPS of <55 %, meaning that the glandular epithelial compartment exceeded that of the stromal compartment within the ROI. The remaining 7/10 cases (blue) do not show image analysis evidence of EIN using the architectural definition of a VPS <55 %. B) All final consensus diagnoses of HwA were correctly found to have stromal areas that exceeded that of the glands by image analysis.

### Expression of p53 and MMR proteins did not discriminate between EIN and HwA

All samples (n=105, HwA (54) plus EIN (51 one sample lost) were successfully processed for p53 staining. All samples, regardless of diagnosis had staining consistent with the presence of a functional ‘wild-type’ protein. There was no evidence of abnormal staining patterns according to the criteria detailed in Kobel et al (Kobel et al., 2016).

All samples were stained for all four MMR proteins (MLH1, MSH2, MSH6 and PMS2). A single patient from the EIN group had focal loss of staining within the EIN region (Supplementary Figure 5). At diagnosis the patient was 51 years old, perimenopausal and presented with heavy menstrual bleeding. The patient was treated surgically with a total abdominal hysterotomy, bilateral salpingo-oophorectomy and peritoneal washings, the final surgical specimens demonstrated a small focus of residual EIN with no evidence of malignancy.

### Loss of ARID1A protein expression is significantly associated with a diagnosis of Endometrial Intraepithelial Neoplasia (EIN)

Loss of ARID1A protein expression was observed in 6/105 (5.7 %) of the EH cases. Samples classified as HwA all contained glands which were immunopositive for ARID1A (Figure 4A). Amongst the EIN samples 1/105 (1.0%) had isolated null glands (Figure. 4C), 4/105 (3.8%) had confluent null glands (Figure. 4D), and 1/105 (1.0%) had complete loss of expression of ARID1A in glands (Figure. 4E). Overall loss of ARID1A protein expression was significantly associated with an EIN diagnosis (Supplementary Table 4, p=0.0112, 2-sided Fisher’s exact test).

**Figure 4.**
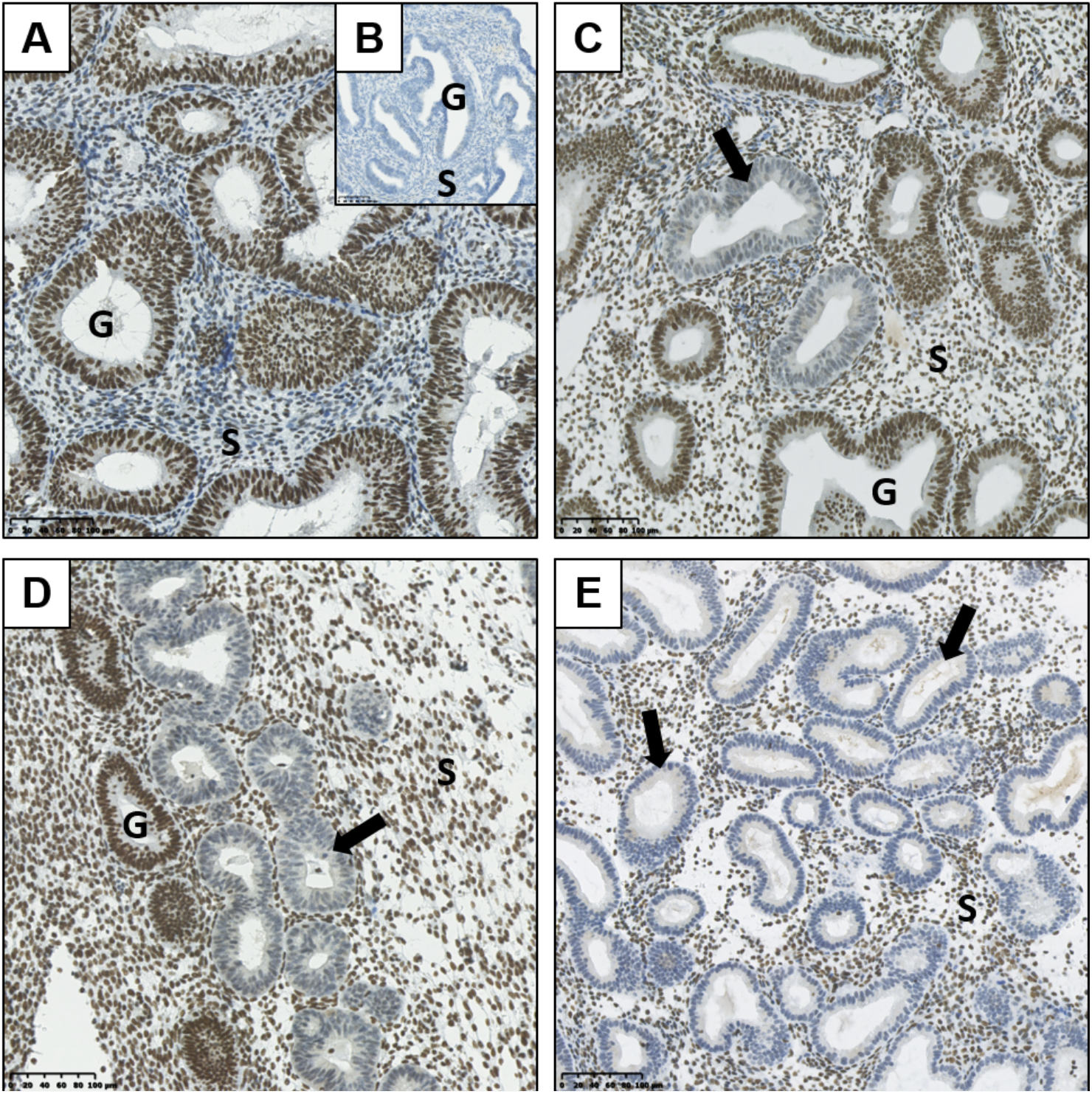
Immunohistochemical expression of ARID1A protein within human endometrial hyperplasia (EH) tissue. A) ARID1A immunohistochemical staining of a tissue section demonstrating hyperplasia without atypia (HwA). Positive ARID1A endometrial glands (G = representative endometrial gland) demonstrated by brown DAB glandular nuclear staining. B) ARID1A negative control. C) Isolated ARID1A null glands (arrow indicating loss of brown DAB glandular nuclear staining) within a tissue section demonstrating Endometrial Intraepithelial Neoplasia (EIN). D) ARID1A confluent null glands (arrow indicating loss of brown DAB glandular nuclear staining) within a tissue section demonstrating Endometrial Intraepithelial Neoplasia (EIN). E) ARID1A complete glandular expression loss (arrow indicating loss of brown DAB glandular nuclear staining) within a tissue section demonstrating Endometrial Intraepithelial Neoplasia (EIN). S = endometrial stromal; used as a positive internal control for ARID1A immunohistochemistry. Varying magnifications – see scale bars.

### Altered HAND2 protein expression is significantly associated with a diagnosis of Endometrial Intraepithelial Neoplasia (EIN)

Altered HAND2 protein expression (either reduced or absent) was observed in 45/103 (43.7%) of the total EH cases examined in this study, with 11/103 (10.7%) demonstrating absent HAND2 expression and 34/103 (33.0%) demonstrating reduced HAND2 expression (Figure 5). The finding of a HAND2 altered immunoexpression pattern was significantly associated with an EH diagnosis of EIN, with 38/49 (77.6 %) of EIN cases demonstrating reduced or absent HAND2 expression compared to 7/54 (13.0 %) of HwA cases (p<0.0001, 2-sided Fisher’s exact test). The finding of absent HAND2 protein expression alone was also significantly associated with an EH diagnosis of EIN. 9/49 (18.4 %) of EIN cases demonstrated absent HAND2 expression compared to 2/54 (3.7 %) of HwA cases (p=0.0235, 2-sided Fisher’s exact test)

**Figure 5.**
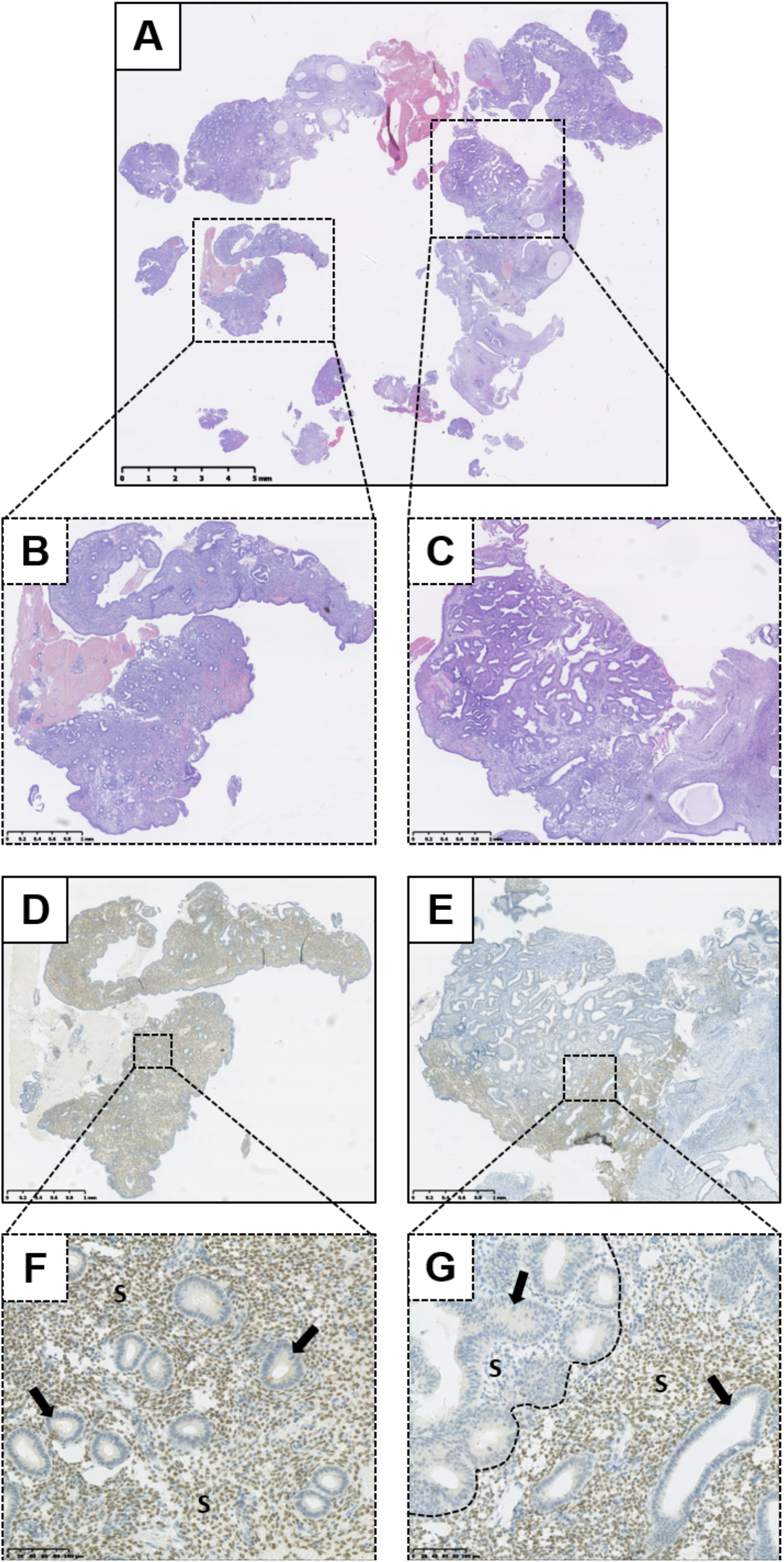
Immunohistochemical expression of HAND2 protein within a tissue biopsy specimen demonstrating Endometrial Intraepithelial Neoplasia (EIN). A) Haematoxylin and Eosin (H&E) stained biopsy specimen containing EIN. B) Higher power representative background endometrium from A. C) Higher power EIN lesion from A. D) HAND2 immunohistochemical staining of B. E) HAND2 immunohistochemical staining of C. F) Higher power image of D demonstrating positive (scored as >50 %) HAND2 stromal nuclear staining within the background endometrium. G) Higher power image of E demonstrating absent (scored as 0 %) HAND2 stromal nuclear staining within an EIN lesion (left of dashed line). Junctional region marked with dashed line. Positive HAND2 (>50 %) staining of background endometrium to right of dashed line. Arrows highlight representative endometrial glands. S= Stroma. Varying magnifications – see scale bars.

### Unsupervised hierarchical cluster analysis (HCA) of PTEN, PAX2 and HAND2 protein expression within EH tissues identified three phenotypes

After a full review of the numbers of samples with changes in patterns of expression of the different proteins cluster analysis was narrowed down to consider only those samples in which immunoscoring results had been obtained for PTEN, PAX2 and HAND2 (n=105 consisting of 51 EIN and 54 HwA based on diagnosis using EIN/WHO2014 criteria; note one sample of EIN was too small for sections to be taken for staining). Examples of staining patterns of PAX2 and PTEN on samples from this dataset have been published previously (Sanderson et al., 2017). Results of the immunohistochemical scoring for these three proteins are summarised in Table 3.

**Table 3.** Immunoscoring results for PTEN, PAX2 and HAND2.

| **Protein** | **EIN, n=51** |  |  | **HwA, n=54** |  |  |
| --- | --- | --- | --- | --- | --- | --- |
|  | *Score 0* | *Score 1* | *Score 2* | *Score 0* | *Score 1* | *Score 2* |
| PTEN (a) | 17 (33.3) | 11 (21.6) | 23 (45.1) | 26 (49.1) | 26 (49.1) | 1 (1.9) |
| PAX2 | 33 (64.7) | 3 (5.9) | 15 (29.4) | 51 (94.4) | 3 (5.5) | 0 |
| HAND2 (b) | 11 (21.5) | 29 (59.2) | 9 (18.4) | 47 (87.0) | 5 (9.3) | 2 (3.7) |

a. one HwA sample stained for PTEN protein was not interpretable. b. two EIN samples stained for HAND2 protein were not interpretable. Percentages in brackets. PTEN - Phosphatase and Tensin Homolog, PAX2 - Paired Box 2 Protein, HAND2 - Heart and Neural Crest Derivatives-expressed 2.

When these results were subjected to unsupervised cluster analysis four subgroups were identified: cluster 1 (n=12), cluster 2 - which was further sub-classified into cluster 2a (n=4) and 2b (n=54) and cluster 3 (n=35) according to dendrogram branch length, which represents the correlation of the scoring data (Figure 6). Clusters 1, 2a and 3 largely contain cases of EIN and could be considered to represent ‘premalignant clusters’ (44/51, 86.3 %), in contrast cluster 2b contained the majority of HwA cases i.e. a ‘benign cluster’ (47/54, 87.0 %): Table 4. The patient demographics for each cluster group are displayed in Supplementary Table 4, demonstrating no significant differences in clinical features between any of the four clusters that could have predicted this grouping.

**Figure 6.**
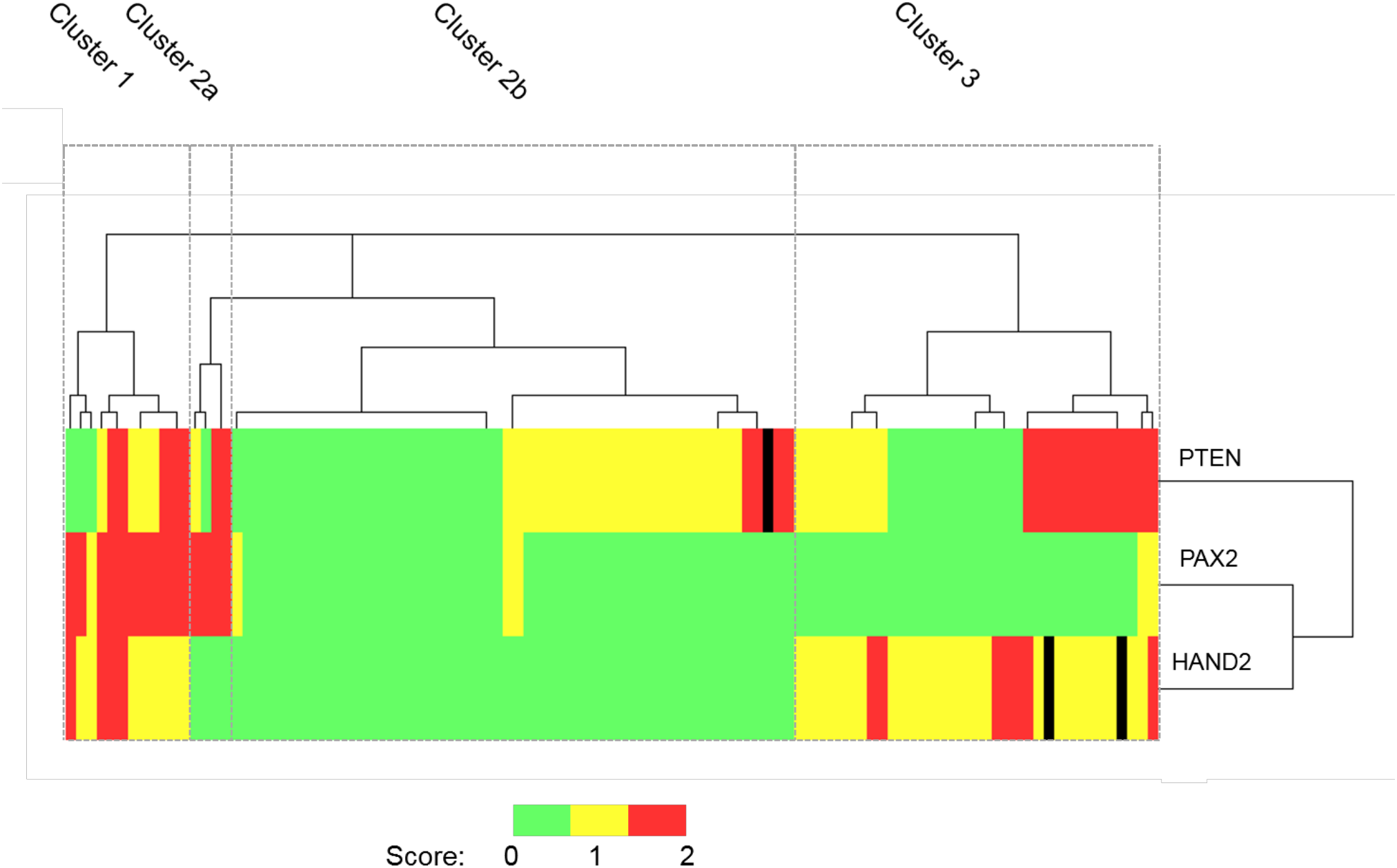
Unsupervised hierarchical cluster analysis of PTEN, PAX2 and HAND2 immunohistochemical staining data. Patient cases orientated along the horizontal axis (n=105). Immunohistochemical markers orientated along the vertical axis. Dendrogram branch length represents the similarity between the results obtained. Heat map colours represent outcome of immunohistochemical scoring as described in M&M. On the basis of the staining patterns EH cases were classified into 4 distinct cluster groups according to the dendrogram: Cluster 1 (n=12), Cluster 2a (n=4), Cluster 2b (n=54) and Cluster 3 (n=35). PTEN - Phosphatase and Tensin Homolog, PAX2 - Paired Box 2 Protein, HAND2 - Heart and Neural Crest Derivatives-expressed 2. Black lines in the heatmap represent cases where the immunoscore was not available.

**Table 4.** Evaluation of each cluster based on classification using WHO and EIN systems.

| **Cluster (%)** | **EIN** | **HwA** | **P value** |
| --- | --- | --- | --- |
|  | N=51 | N=54 |  |
| 1 | 12 (23.5) | 0 | 0.0001 |
| 2a | 4 (7.8) | 0 | 0.057 (ns) |
| 2b | 7 (13.7) | 47 (87) | <0.0001 |
| 3 | 28 (54.9) | 7 (11.3) | <0.0001 |

EIN = Endometrial Intraepithelial Neoplasia. HwA = Hyperplasia without atypia. Percentages
in brackets. Statistical analysis using a Fisher's exact test, 2 sided. \*\*\*p<0.001, \*\*\*\*p<0.0001, ns = non-significant.

### Progression of EH to endometrioid endometrial cancer

The immunophenotype as determined using detection of PTEN, PAX2 and HAND2 proteins, for each case of EH that subsequently progressed to EC was also determined. Importantly, n=9 (90 %) of the EH that progressed to EC demonstrated reduced expression of HAND2 protein. PAX2 protein expression varied dependent on cluster grouping (altered PAX2 expression was exclusive to cluster 1 EH cases), with n=3 (30 %) of the overall EH cases that progressed to EC demonstrating a change in PAX2 protein. PTEN protein loss was found in n=7 (70 %) of the EH cases which progressed to EC, incorporating n = 2 (20 %) cases which exhibited isolated PTEN null glands, and n=5 (50 %) which exhibited a PTEN null region.

Although p53 expression was not altered ARID1A protein expression was lost in n=2 (20%) of the EH cases that progressed to EC. Both cases of ARID1A loss in EH were associated with an EC that was likely concurrent (diagnosed within 12 months of initial EH biopsy). Examples of the phenotype of patients in cluster 1 that progressed to EC are shown in Supplementary Figure 6 and three of those from Cluster 3 are illustrated in Figure 7.

**Figure 7.**
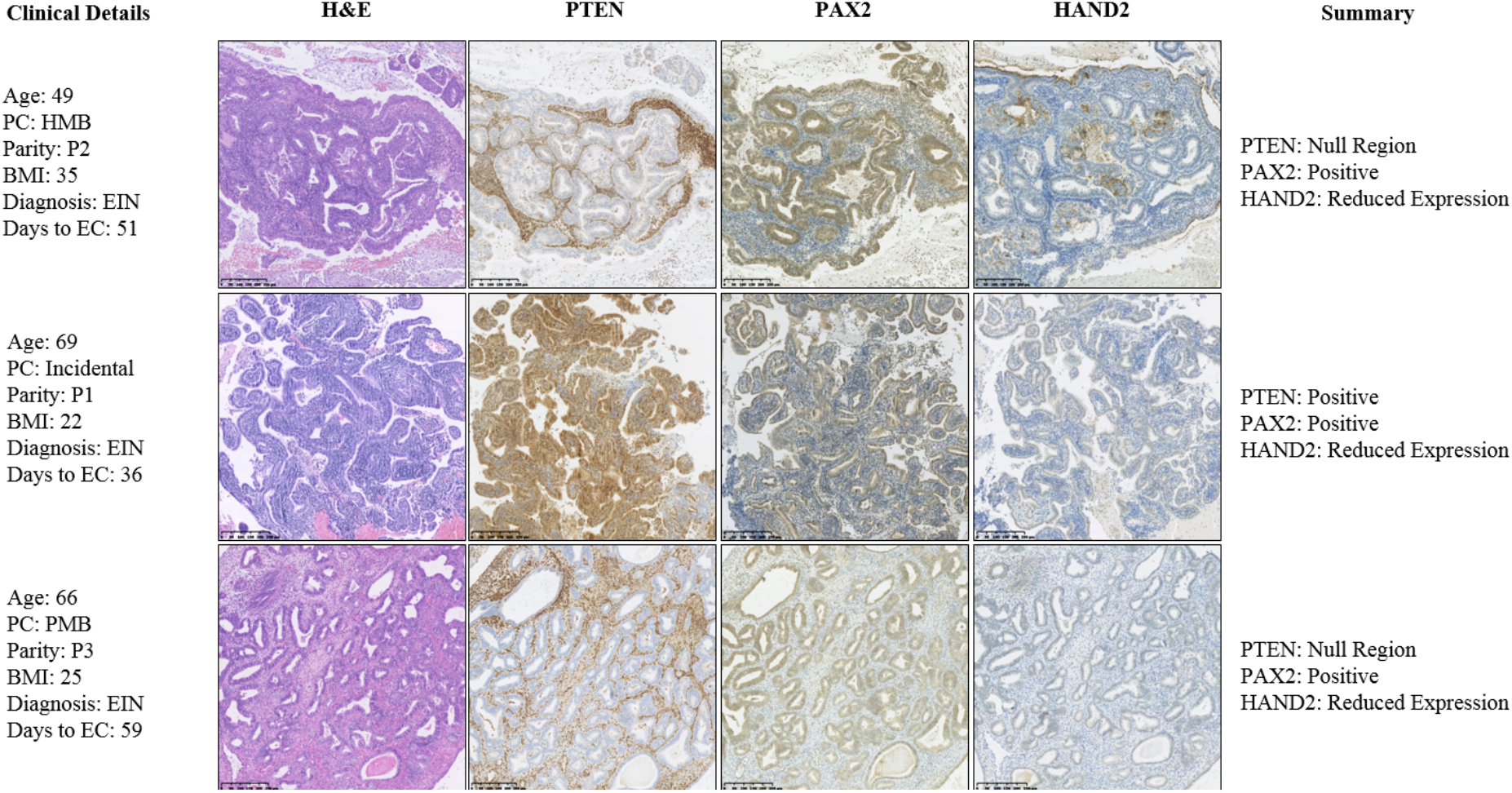
Immunohistochemical phenotype of Cluster 3 cases that progressed to malignancy based on biopsy at time of diagnosis of EH. 6 patients from cluster 1 had a subsequent endometrioid endometrial cancer diagnosis. Images from three patients are used to illustrate the expression patterns of the PTEN, PAX2 and HAND2 for each with pertinent clinical details. The selected images for each case represent serial tissue sections of the most abnormal hyperplastic region. EC - Endometrial cancer, PMB - Postmenopausal bleeding, HMB - Heavy menstrual bleeding. Varying magnifications – see scale bar

## Discussion

Rates of endometrial cancer in the UK have increased by 56% since the 1990s [https://www.cancerresearchuk.org/]. Although often considered a cancer of post-menopausal women a marked increase has also been noted in younger women with a 44% increase in incidence in 25- to 49-year-olds between 1993 and 2018. One of the key risk factors associated with increasing rates of both pre-malignant and malignant changes in the endometrium is obesity with meta-analysis suggesting that the risk of EC is 81% higher per 5-unit BMI gained during adulthood (Stevens et al., 2014). Other factors including longer menstrual lifespans, diabetes mellitus and polycystic ovarian syndrome (PCOS) or genetic factors such as Lynch syndrome are all also considered risk factors for both endometrial cancer and endometrial hyperplasia (Sanderson et al., 2017). A priority setting partnership, that brought together patients as well as health care professionals agreed one of the top-ten unanswered questions for EC research, was for development of a personalised risk score for developing EC (Wan et al., 2016). This is of particularly relevance to younger women many who would prefer to avoid hysterectomy so they can have a future pregnancy.

In this study we capitalized on an extensive local tissue biobank set up in 2015 with ethical approval that enabled us to access archival material held in the histopathology diagnostic archive. Following a search of the database associated with this archive we identified 145 tissue samples originally classified using the WHO94 criteria as having a histology consistent with endometrial hyperplasia between 2004 and 2009. Our team also included 3 pathologists with extensive experience of gynaeological pathology that afforded us a unique opportunity to conduct a fresh evaluation of the original biopsy based on the original WHO94 criteria and to compare results obtained using the revised criteria published in 2014. The latter simplified the diagnostic categories and incorporated an evaluation of stromal to epithelial ratio and gland crowding considered characteristic of premalignant EIN lesions (Mutter, 2002). In agreement with other reports we found a marked improvement in agreement between pathologists using the 2014 criteria although it has been suggested further studies are still needed to determine best methods with regard to coexistent cancers (Raffone et al., 2019, Travaglino et al., 2019).

Ultimately the purpose of any EH pathological classification is to identify women who are at a higher risk of progression to EC so that a care pathway can be agreed and implmented. Historically a 1985 study by Kurman *et al*. is frequently cited that appears to be the basis for the widely held opinion that approximately one third of patients with complex atypical hyperplasia (CAH) will eventually develop EC if they do not undergo hysterectomy (Kurman et al., 1985). Baak *et al*. claimed that the newer EIN classification system could more accurately predict progression to EC than the WHO94 system (Baak et al., 2005a) but others had reported that both EIN and atypical hyperplasia have similar risks of progression to EC when followed-up for 12 months after the index diagnosis (Lacey et al., 2008a). In the current study we examined all the available clinical records to see whether women who originally had a diagnosis of EH based on the WHO94 criteria (n=118, from an original sample of n=125 owing to losses to follow-up) to see if a subsequent EC was detected. In this same sample set we also had data from re-evaluation of the original histology using the updated EIN/WHO2014 criteria allowing a comparison to be made. Numbers were limited because many of the women given an original diagnosis of CAH were treated surgically. Within our cohort 80% (8/10) cases of EC were diagnosed within 12 months of the index biopsy being re-classified as EIN. Sensitivity and specificity data from this study were in keeping with that described by others (Baak et al., 2005a) and we found that the WHO94 system was not as good at predicting the absence of subsequent EC (Negative predictive value, NPV, of 91.6 %) when compared to the EIN/WHO2014 system (NPV 98.4%).

With advances in whole-slide scanning technologies and digital imaging becoming more mainstream there is a rapidly growing field of digital pathology (Jahn et al., 2020). Increased online sharing of information between experts particularly for complex cases is also encouraged as there is a reported national and global shortage of pathologists. In the current study we used a digital tool kit to evaluate whether the data generated by an automated analysis of the stromal volume could increase the reliability of the diagnosis of EIN. In our dataset we had 21 cases considered suitable for this form of evaluation and the method applied showed good agreement with a diagnosis of HwA but suggested the 55% cutoff for EIN might need to be reconsidered. Further studies using larger numbers of samples and other digital platforms are recommended before the computerized analysis is widely applied but we believe this kind of methodology should be more widely rolled out.

Whilst a number of changes in expression of proteins in the endometrium have been investigated for their links with progression to malignancy to date no single candidate has reliably and reproducibly been shown to predict malignant progression although a number have shown promise (Sanderson et al., 2017). In this study we investigated a number of these candidates to see how they aligned with the diagnostic criteria based on H&E and also grouped data using unsupervised clustering to see if combinations of markers might increase predictive power and improve decision making. Consistent with reports from other studies we noted changes in expression of both PTEN (Monte et al., 2010, Mutter et al., 2000) and PAX2 (Quick et al., 2012). in agreement with reports by Mutter et al that loss of PTEN was not a good predictor or progression to EC (Lacey et al., 2008b) we did not find loss of PTEN was a good predictor of whether samples were classified as HwA or EIN. We only found one sample with changes in MMR proteins and none with altered p53. The latter appears in agreement with a recent paper which re-assed p53 staining in ∼200 endometrial cancers and reported abnormal staining in only 14.5% of stage 1A samples (Kang et al., 2022). A previous study identified HAND2 as a candidate for epigenetic deregulation in EC (Jones et al., 2013) although another study failed to find evidence of change that could distinguish hyperplasia with or without atypia (Buell-Gutbrod et al., 2015) they did think it might be a useful biomarker. In this study we found the combination of scoring for HAND2, PTEN and PAX2 was able to align staining patterns with diagnosis of EIN or HwA based on diagnostic criteria and might be useful in identifying those most likely to have benign disease. It will be interesting to see these 3 markers also yield the same results in other datasets or prospective sample collections.

With cases of EH and cancer rising women there is an increased emphasis on non-invasive methods of that can reduce costs of initial screening without the need for the extensive evaluation of tissue samples as described above. A recent review by authors based at the Mayo Clinic in the USA highlighted the need for new approaches highlighting the highly variable and high cost of standard investigations (2-25K USD) associated with an investigation prompted by presentation with a diagnosis of abnormal or post-menopausal bleeding even when the results ultimately suggested no further intervention was required (Warring et al., 2022). Imaging of the endometrium, genetic diagnosis of at-risk subgroups, or blood tests based on putative biomarkers are all under active investigation. For example, a systematic review of data on whether endometrial thickness is a useful tool for evaluating symptoms such as bleeding in post-menopausal women concluded that the incidence of endometrial carcinoma, hyperplasia or polyps was significantly higher if the thickness was >5mm and that this could be useful to identify those at risk and therefore merited further investigation (Su et al., 2021).

The emergence of a genetic profile for EC which stratified some patients as low risk (Kandoth et al., 2013) has increased in screening patients with EP for genetic risk factors. However recent reports that cancer associated mutations also occur with high frequency in normal endometrium have however led to calls for some caution in interpretation of general screening for early detection of cancer and the need to consider not just one but multiple genetic hits as pathogenic (Kyo et al., 2020). One group who are candidates for genetic screening are women from families with Lynch syndrome, a condition associated with higher rates of colorectal cancer as a result of DNA mismatch repair deficiency (Poulogiannis et al., 2010). Lynch syndrome is also associated with an increased risk of developing EC and ovarian carcinoma. A systematic review of the literature suggested 3% of EC could be attributed to Lynch syndrome (Ryan et al., 2019) with loss of MLH1 due to promoter hypermethylation being one cause (Ryan et al., 2020). In our study we screened for MMR deficiency as a potential cause of EIN and identified one patient from 51 diagnosed with EIN where the loss of expression was in the abnormal area of tissue (∼2%) which was lower than we expected but probably reflected the unselected nature of our patient sample group.

Proteomic methods such as mass spectrometry have also been deployed in efforts to develop a non-invasive blood test for EC. A large number of candidates have been proposed including hormones, cancer associated antigens such as CA125, enzymes, enzyme inhibitors and growth factors as recently reviewed in Njoku et al (Njoku et al., 2019). Serum HE4 appears overexpressed in patients with EC and has shown promise for predicting response to progestin therapy (Behrouzi et al., 2021, Behrouzi et al., 2020). A combination of CA124, HE4 and clinical characteristics such as BMI has been reported to have a specificity of ∼85% with serum HE4 predicting deep tissue invasion (Knific et al., 2017). More recent studies have endorsed these markers as useful in diagnosis of recurrence and metastases (O’Toole et al., 2021, Quan et al., 2021). Whilst none of the biomarkers are routinely used in clinical practice to diagnose endometrial hyperplasia/EIN it is notable that reductions in circulating biomarkers of insulin resistance and inflammation were detected in women who underwent bariatric surgery which resulted in reversal of neoplastic changes including atypical hyperplasia and EC (MacKintosh et al., 2019) consistent with the suggestion that more attention should be paid to immune surveillance in EC prevention (Naqvi et al., 2021).

### Summary and conclusions

Rates of EC are rising and early and accurate diagnosis of precursor lesions in endometrium is an important challenge for the gynaecologist and pathologist particularly when women are pre-menopausal and may wish to avoid hysterectomy. We have found application of the revised EIN/WHO2014 criteria is more likely to give a consensus diagnosis and that computer aided imaging of gland to stroma ratio is a useful adjunct to assist diagnostic accuracy. We propose that stratification of risk of malignant progression could also be improved by using a combined immunostaining score based on evaluation of HAND2, PAX2 and PTEN. Further larger prospective studies using these criteria as well as other tools including measurement of blood biomarkers are recommended.

## Supporting information

Sanderson_Suppl Figs and Tables

## Data Availability

All data produced in the present work are contained in the manuscript

