## Supplementary material for "Improving the diagnosis of endometrial hyperplasia using computerized analysis and immunohistochemical biomarkers": Sanderson_Suppl Figs and Tables

**Supplementary Figure 1. Application to the SE Scotland HSS Bioresource**

NHS Lothian

CHIEF SCIENTIST OFFICE

SR465

### REQUEST FOR ACCESS TO SOUTH EAST SCOTLAND HSS

### (formerly SAHSC) BIORESOURCE

This application form is for the purpose of making a request to collect, use and process tissue/data via the HSS (SAHSC) BioResource. The tissue/data must be utilised within the laboratory and by personnel that fall under the supervision of the Principal Investigator listed on the application. Any transfer of tissue/data to personnel or laboratories that are not under the supervision of the indicated PI requires the following:

- An explanation of the need to transfer the tissue/data and benefit to the investigator's research
- A copy of the enclosed BioResource agreement form signed by the collaborator

The South East Scotland HSS (SAHSC) BioResource does not supply tissue/data solely for distribution to third party researchers, those researchers should apply to the BioResource directly.

The requested information is necessary in order to document your request for permission to collect, or access tissue/data and to ensure that the South East Scotland HSS (SAHSC) BioResource operates within the guidelines of the Tissue Act Scotland, 2006.

When submitting a written request for permission to collect or access tissue/data:

- Please print neatly or type.
- Patient identity is confidential. Samples will be coded and supplied with a minimum data set.
- Researchers are required to cover the cost of transport of their samples and supply appropriate customs declarations if applicable.
- For additional information please contact the South East Scotland HSS (SAHSC) BioResource at, or
- Please send completed request forms and proof of ethical approval where applicable to Frances Rae with a copy to Craig Marshall at the above e-mail address.
- Application will then be sent to the appropriate Scientific Review Committee for consideration and approval. No tissue or data will be released prior to approval being granted.

**REQUEST FORM.**

| | |
|---|---|
| Name & Address of PI:<br><br><br><br><br><br><br>e-mail address: | Professor Philippa Saunders<br>Professor of Reproductive Steroids,<br>MRC Centre for Inflammation Research,<br>The Queen's Medical Research Institute,<br>47 Little France Crescent<br>Edinburgh<br>EH16 4TJ<br>0131-242-6388 (office)<br><br> |
| Study Title:<br><br>Ethical Status: | Early Diagnosis and Treatment Strategies for Endometrial Cancer<br><br>N/A |
| Sponsor: | N/A |
| Funding Body: | Cancer Research UK / Edinburgh Cancer Research Centre |
| Material/Data Requested:<br><br>Please be specific as to the nature of the material/data and indicate exactly the type and quantity eg fresh frozen, paraffin embedded, archival or prospective etc.<br><br>If the request is for sections, please specify quantity, thickness of sections required, and type of slide eg plain or adhesive. | We would like to request endometrial tissue samples and have access to anonymised patient medical history data stored within NHS Lothian and the Department of Pathology at the Royal Infirmary of Edinburgh.<br><br>Via our NHS Lothian/Clinical collaborator – Professor Alistair Williams, Chair of Gynaecological Pathology, a patient cohort of interest has been identified. These patients underwent endometrial sampling and/or surgery between the years 2004-2009 and were diagnosed with the condition endometrial hyperplasia (Now also referred to as Endometrial Intraepithelial Neoplasia – EIN).<br><br>From this patient cohort we would like to obtain the original index diagnostic tissue sample(s), any prior / subsequent tissue samples taken in the diagnostic/treatment timeline and also anonymised demographic/medical history information about these patients from their medical records (paper or electronic.)<br><br>These data/tissue sections will be used within the University of Edinburgh for further research into the classification and understanding of the condition endometrial hyperplasia/EIN by examining expression of putative biomarkers of the condition. |

| | |
|---|---|
| | This will involve immunohistochemical staining for proteins previously identified as diagnostic of 1) the proliferation status of cells within the tissue; 2) response to steroids [e.g. receptor proteins] 3) epithelial-mesenchymal transition 4) mismatch repair activity 5) inflammation. Information from this study will be analysed and correlated along with the medical history data and patient demographics.<br><br>If unstained slides are provided, we would like the archival tissue to be from FFPE blocks, cut at 5 microns and on adhesive glass slides.<br><br>There are 143 patients within our identified cohort and we would initially require 50 unstained sections from the index diagnostic biopsy of each patient.<br><br>From a subset of the cohort (unknown quantity at present) we would also want 50 unstained sections from selected pre and post index biopsy samples along the diagnostic timeline in order to map disease progression.<br><br>Medical history and demographic data would include: age, BMI, parity, hormone use, family history, past medical history, past surgical history, smoking status and other relevant risk factor(s) data for development of endometrial hyperplasia/EIN. |
| Synopsis of project (100 – 200 words approximately): | Endometrial cancer is a common malignancy of the uterine lining; rates are rising and there is an increasing incidence of pre-menopausal disease which is more difficult to diagnose.<br>Sex-steroids (especially unopposed oestrogens) have been shown to play a crucial role in the development of endometrial cancer, with lifetime exposure influencing cell proliferation in normal and malignant endometrial tissue. A recent integrated genomic and proteomic analysis has provided new insights into the biology and classification of endometrial cancer.<br>This project will investigate endometrial pre-malignant neoplasia using a range of methods in order to develop improved strategies for earlier diagnosis and treatment.<br>In 1994 The World Health Organisation (WHO) classified the condition known as 'Endometrial Hyperplasia'. The WHO classification of endometrial hyperplasia consists of four categories, of which complex hyperplasia with nuclear atypia is considered to have the greatest risk of progression to endometrioid endometrial carcinoma. However, there is often considerable variability amongst pathologists when diagnosing specimens using the WHO nomenclature<br>A new classification of the endometrial intraepithelial neoplasia (EIN) was published in 2000. It has been claimed that the EIN system better predicts disease progression.<br><br>We wish to investigate the following: |

| | |
|---|---|
| | (i) What are the key cellular changes that characterise these precursor lesions of endometrioid endometrial carcinomas?<br><br>(ii) What are the patterns of expression of steroid receptors and steroid responsive genes in the precursor lesions of endometrioid endometrial carcinomas and can they predict responsiveness to therapies designed to interrupt the precursor-cancer transition? |
| If tissue collection is prospective, estimated number of patients to be consented. | N/A |
| Material disposal or storage plan: | 1) Slides would be physically stored within the University of Edinburgh, MRC Centre for Inflammation Research and if no longer required, sensitive disposal would be undertaken in line with the University of Edinburgh guidelines on disposal of human tissue waste.<br><br>2) Electronically stored (Slides scanned) on secure server as a future reference library. |
| Date Required: | ASAP |
| Shipping Address<br>(if different to above): | Saunders Lab, West-Block<br>MRC Centre for Inflammation Research,<br>The Queen's Medical Research Institute,<br>47 Little France Crescent<br>Edinburgh, EH16 4TJ |

**AGREEMENT FOR USE OF TISSUE/DATA.**

The recipient agrees that any tissue/data provided by the South East Scotland HSS (SAHSC) BioResource will only be used for the purposes specified in this application and will only be used for the common good in scientific research or education. The recipient shall maintain retrievable records linking the material and accompanying data to the terms of acquisition. The recipient agrees not to attempt to obtain information identifying the individuals donating tissues/data to the South East Scotland HSS (SAHSC) BioResource. The recipient agrees they shall not sell any portion of the tissues/data provided by the South East Scotland HSS (SAHSC) BioResource, or products directly extracted from tissue material (e.g. protein, mRNA or DNA). The recipient also agrees that they shall not transfer tissue/data (or any portion thereof) supplied by the South East Scotland HSS (SAHSC) BioResource to third parties without prior written permission of the South East Scotland HSS (SAHSC) BioResource. Any subsequent transfer that may be made to other parties, with prior agreement from the South East Scotland HSS (SAHSC) BioResource, will require signature of this agreement between the final recipients of the material and the South East Scotland HSS (SAHSC) BioResource.

The recipient understands that while the South East Scotland HSS (SAHSC) BioResource attempts to avoid providing tissues that are contaminated with highly infectious agents eg hepatitis or HIV, all tissues should be handled as if potentially infectious. The individuals who have supplied tissue to the South East Scotland HSS

(SAHSC) BioResource have not agreed to have clinical tests performed on this tissue, therefore, the recipient agrees not to perform such tests on the tissues supplied by the South East Scotland HSS (SAHSC) BioResource. The recipient acknowledges that the institution where the tissue/data will be used follows the relevant Human Tissue Authority or appropriate local regulations for handling human specimens and will instruct their staff to abide by those rules. The recipient further agrees to assume all responsibility for informing and training personnel in the potential risks and safety procedures for the handling of human tissues.

Tissues are provided as a service to the research community without warranty of merchantability or fitness for a particular purpose or any other warranty, express or implied. The South East Scotland HSS (SAHSC) BioResource accepts no responsibility for any injury (including death), damages or loss that may arise either directly or indirectly from their use.

The recipient agrees to acknowledge the contributions of the South East Scotland HSS (SAHSC) BioResource in all publications resulting from the use of these tissues/data

The institution agrees to assume all risks and responsibility in connection with receipt, handling, storage and use of tissue/data from the South East Scotland HSS (SAHSC) BioResource. It further agrees to indemnify and hold harmless the South East Scotland HSS (SAHSC) BioResource from any claim costs, damages or expenses resulting from the use of the tissue/data provided by the South East Scotland HSS (SAHSC) BioResource. The undersigned warrant that they have authority to execute this agreement on behalf of the recipient institution.

By my signature I agree to the terms set out in the above agreement

| | |
|---|---|
| Name of Principal Investigator: | Professor Philippa TK Saunders |
| Institution: | The University of Edinburgh |
| Signature: | |

### Supplementary Figure 2. Approval of sample requests from HSS Bioresource

| Document Name | QF-TGU-A-SAMREQA VERSION 1.0 | Page | 1 of 1 | Review date | 15-Jul-2016 |
|---|---|---|---|---|---|

#### NHS LOTHIAN SAHSC BIORESOURCE SAMPLE REQUEST ANSWER FORM

| | |
|---|---|
| Sample Request number: | SR465 |
| Name of Researcher: | Professor Philippa Saunders |
| Address of Researcher: | Director, Postgraduate Research CMVM<br>MRC CIR<br>QMRI<br>Edinburgh |
| Study Title: | Early Diagnosis and Treatment Strategies for Endometrial Cancer |
| Ethical status: | 13/ES/0126 |
| Material Requested | Release and use of the followng is approved in principle for the purposes of this project:, but we will be unable to meet the requirement for 50 sections from the blocks.<br><br>Anonymised archival endometrial tissue samples and linked patient medical history data as described on the request form.<br><br>Paraffin sections will be supplied, but the number of slides will be significantly less than 50, and further discussion with Prof Williams will be necessary. |

**REQUEST AUTHORISED**

| | |
|---|---|
| Date: | 12-Feb-2015 |
| Authorised by: | Frances Rae |

**REQUEST REJECTED**

| |
|---|
| Date: |
| Authorised by: |
| Reason |

| | |
|---|---|
| **Author : Frances Rae** | **Date : 15-Jul-2010** |
| **Authority for Issue : Craig Marshall** | **Date : 15-Jul-2010** |
| **Quality Checked : Craig Marshall** | **Date : 15-Jul-2010** |

**Supplementary Figure 3. Proforma used by pathologists for independent assessment of the n=125 tissue samples.**

| | | | | |
|---|---|---|---|---|
| **Sample number:**<br>**Date reviewed:**<br>**Reviewer:** | | | | |
| Adequate endometrial tissue for diagnosis? | **YES** | **NO** | **Only if NO: Tick all that apply** | |
| | □ | □ | Inadequate Quality | □ |
| | | | Processing / technical problems | □ |
| | | | Other: Specify | |
| **Diagnosis:** | | | | |
| Benign Endometrium | **YES** | **NO** | **Only if YES: only tick one** | |
| | □ | □ | Atrophic | □ |
| | | | Inactive | □ |
| | | | Proliferative | □ |
| | | | Disordered Proliferative | □ |
| | | | Secretory (including progestin & OCP effect) | □ |
| | | | Menstrual | □ |
| | | | Endometritis | □ |
| | | | Other: Specify | |
| Endometrial Hyperplasia | □ | **WHO 2014** | Hyperplasia without Atypia | □ |
| | | | Endometrioid intraepithelial neoplasia (EIN) | □ |
| | □ | **WHO 1994** | Simple, non-atypical hyperplasia | □ |
| | | | Complex non-atypical hyperplasia | □ |
| | | | Simple atypical hyperplasia | □ |
| | | | Complex atypical hyperplasia | □ |
| Malignant Neoplasm | □ | □ | Endometrial neoplasm | □ |
| | | | Type: Specify | |
| | | | Other malignant neoplasm | □ |
| | | | Type: Specify | |
| **Polyp:** | | | | |
| Endometrial polyp | **YES** | **NO** | **Only if YES: only tick one** | |
| | □ | □ | Atrophic | □ |
| | | | Functional | □ |
| | | | Hyperplastic | □ |
| **Notes:** | | | | |

**Supplementary Figure 4 A representative example of layered segmentation analysis of a section of endometrial tissue containing a region of hyperplasia analysed using the TissueGnostics 'H&E app'.**

Sections of endometrial tissue biopsises stained with H&E were converted into digital images using a NanoZoomer-XR scanner in 40x mode and stored as NanoZoomer Digital pathology files (.pdpi) which were imported and processed using StrataQuest analysis software. Individual regions of interest (ROI) were identified and analysed using the software to generate a numerical area/volume analysis of different compartments within the tissue. In the image supplied the following colour codes are used: dark green = endometrial stroma; blue = endometrial gland connecting with a lumen; red = endometrial gland without connection to lumen/surface; light green = lumen of a gland.

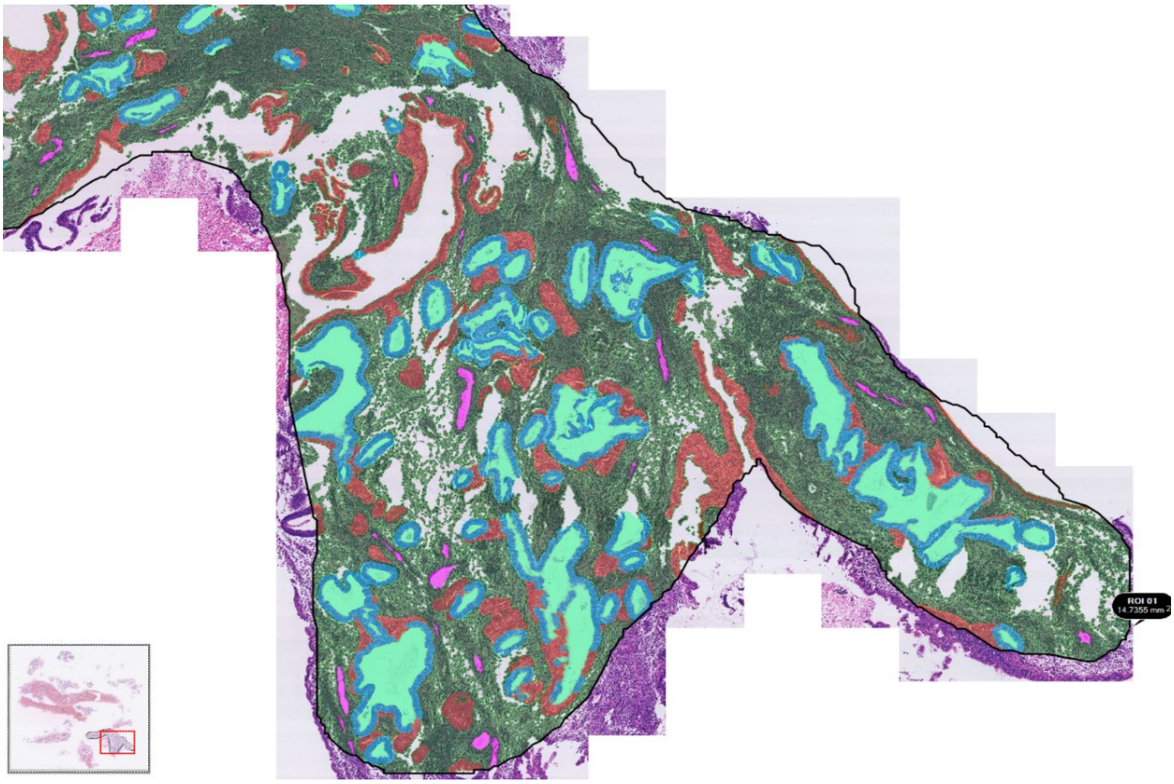

**Supplementary Figure 5. Deficient MMR in a region of tissue diagnosed as EIN**

A) Low power H&E image of an EH lesion containing a focus of EIN (box B), normal endometrium (box C). B) EIN lesion: C) Endometrial glands/normal background tissue. D) Higher power view of the EIN lesion from B showing crowding of glands. B1) Loss of MLH1 expression in the glandular nuclei of the EIN lesion (loss of nuclear brown DAB staining within the glandular nuclei). B2) Normal expression of MSH2 in the same sample. B3) Normal expression of MSH6 in the same sample. B4) Loss of binding partner PMS2 expression in the glandular nuclei of the same sample. D1/ D4) HP images demonstrating loss of MLH1 and PMS2 expression respectively (arrowed), background endometrium positively stained and included in C1/C4 for comparison. D2/D3) HP MSH2 and MSH6 positive expression, background endometrium positively stained and included in C2/C3 for comparison.

G = endometrial gland, S = stroma. Varying magnifications – see scale bars.

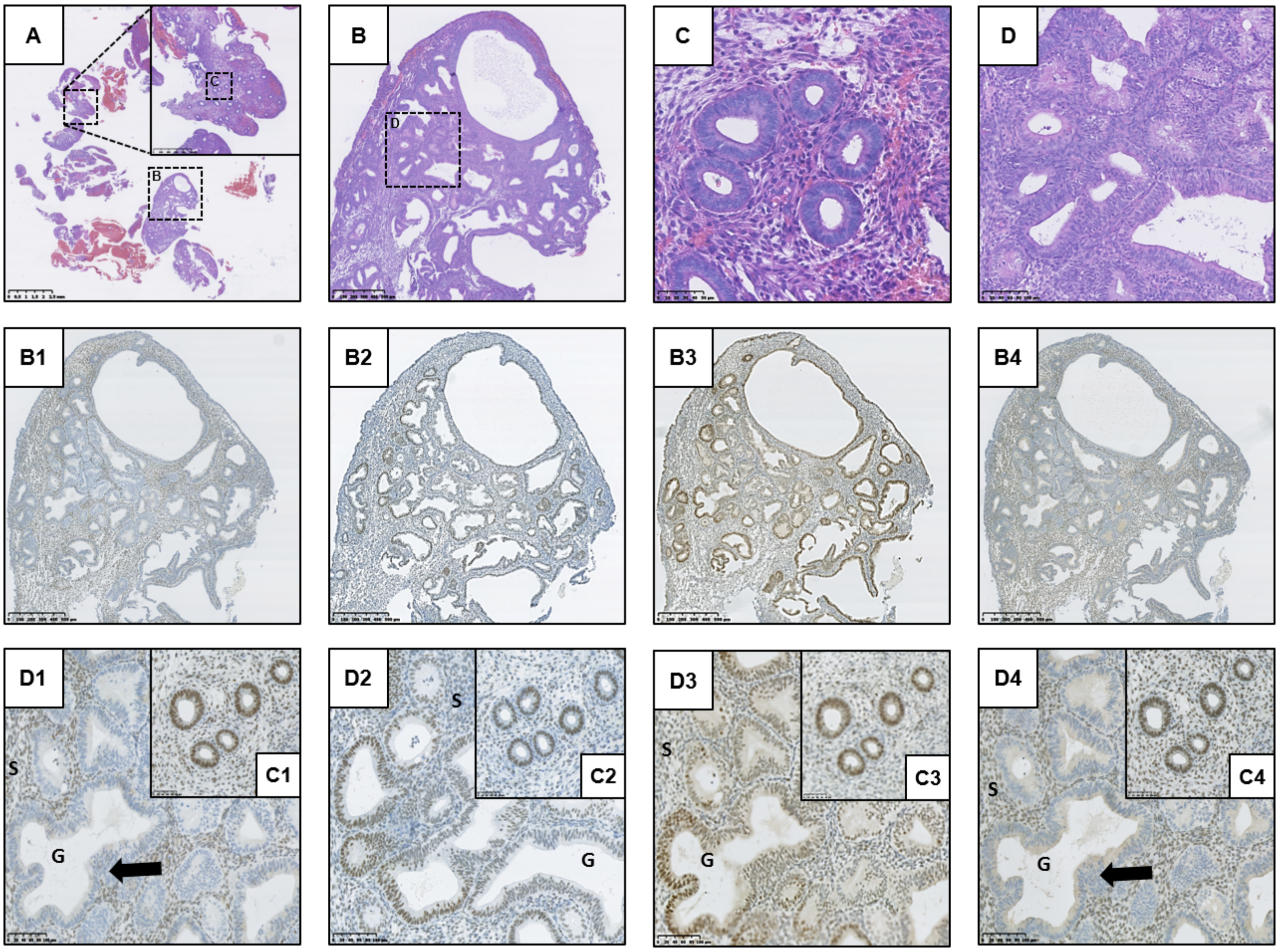

**Supplementary Figure 6. Immunohistochemical phenotype of Cluster 1 EH cases that progressed to malignancy.**

This analysis focused on the n=3 patients from cluster 1 that had a subsequent confirmed diagnosis of endometrioid endometrial cancer. The image provides a summary of the relevant clinical details, illustrates the appearance of the sample at time of EH diagnosis and the expression patterns of PTEN, PAX2 and HAND2 proteins. The selected images for each case represent serial tissue sections of the most abnormal hyperplastic region.

Key H&E - Haematoxylin and Eosin; PTEN - Phosphatase and Tensin Homolog; PAX2 - Paired Box 2 Protein; HAND2 - Heart and Neural Crest Derivatives-expressed 2.

EC - Endometrial cancer, PMB - Postmenopausal bleeding, HMB - Heavy menstrual bleeding, IMB - Intermenstrual bleeding. Magnifications vary – scale bars are shown.

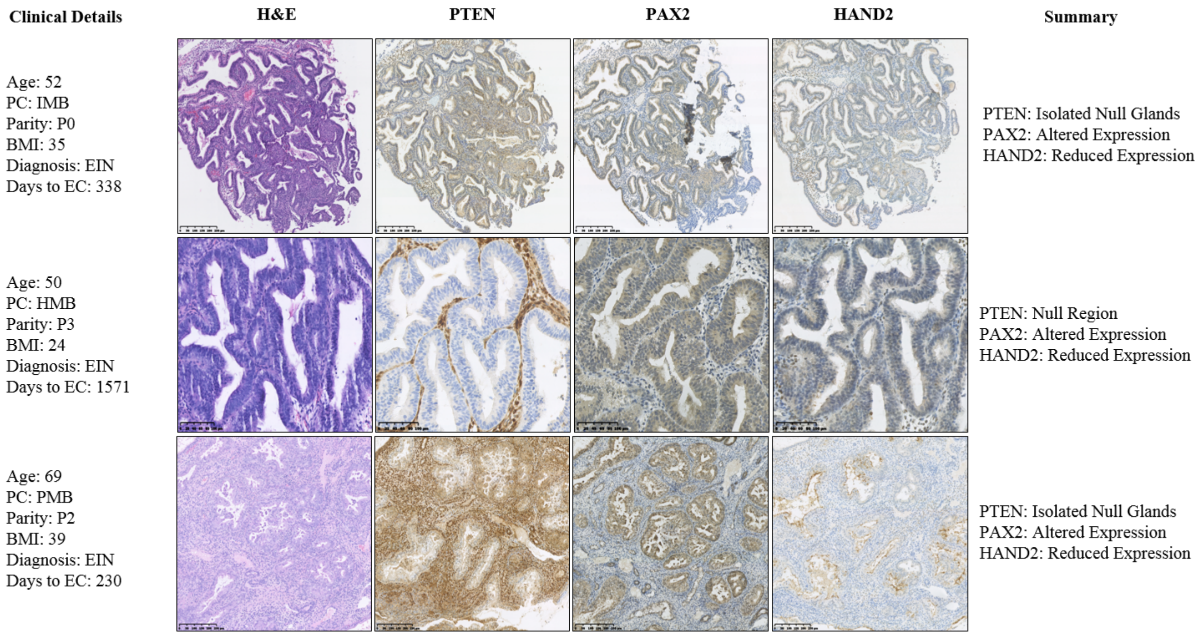

SUPPLEMENTAL TABLES

**Supplemental Table 1:** Primary antibodies and detection systems used for chromogenic immunohistochemistry

| Antigen | Species | Supplier | Cat. No | Dilution | ImmPRESS™ polymer |
|---|---|---|---|---|---|
| Anti-PTEN (clone 6H2.1)* | Mouse (Monoclonal) | Agilent Dako | M362729-2 | 1:300 | ImmPRESS™ Anti-Mouse (Vector MP 7402) - DAB |
| Anti-PAX2 (clone Z-RX2) | Rabbit (Polyclonal) | Invitrogen | 71-6000 | 1:900 | ImmPRESS™ Anti-Rabbit (Vector MP 7401) - DAB |
| Anti-ARID1A* | Rabbit (Polyclonal) | Sigma-Aldrich | HPA005456 | 1:2000 | ImmPRESS™ Anti-Rabbit (Vector MP 7401) - DAB |
| dHAND (clone M-19) | Goat (Polyclonal) | Santa-Cruz | sc-9409 | 1:250 | ImmPRESS™ Anti-Goat (Vector MP 7405) – DAB |
| p53 (clone DO1) | Mouse (Monoclonal) | Santa-Cruz | sc-126 | 1:500 | ImmPRESS™ Anti-Mouse (Vector MP 7402) - DAB |
| MLH1 (clone C-20) | Rabbit (Polyclonal) | Santa-Cruz | sc-582 | 1:100 | ImmPRESS™ Anti-Rabbit (Vector MP 7401) - DAB |
| MSH2 (clone FE11) | Mouse (Monoclonal) | Millipore Merck | MABE284 | 1:500 | ImmPRESS™ Anti-Mouse (Vector MP 7402) - DAB |
| MSH6 (clone 44) | Mouse (Monoclonal) | BD Biosciences | 610919 | 1:250 | ImmPRESS™ Anti-Mouse (Vector MP 7402) - DAB |
| PMS2 (clone A16-4) | Mouse (Monoclonal) | BD Pharmingen | 556417 | 1:300 | ImmPRESS™ Anti-Mouse (Vector MP 7402) - DAB |

*Internal positive control present within endometrial tissues.

**Supplementary Table 2. Subgroup breakdown recorded within the original index cases in the category of complex atypical hyperplasia (CAH)**

| Subgroup within CAH | number | percentage |
|---|---|---|
| Complex atypical hyperplasia | 10 | 41.6 |
| Complex hyperplasia with mild atypia | 6 | 24 |
| Complex hyperplasia with moderate atypia | 4 | 16 |
| Complex hyperplasia with severe atypia | 2 | 8 |
| Severe atypical hyperplasia | 1 | 4 |
| Complex hyperplasia with minor atypia | 1 | 4 |
| Total | 24 | |

**Supplementary Table 3: Demographics and clinical features of patients with a diagnosis of endometrial intraepithelial neoplasia (EIN) or hyperplasia without atypia (HwA) based on a consensus diagnosis by two independent gynaecological pathologists.**

| | *EIN n=52 (%)* | *HwA n=54 (%)* | *P Value* |
|---|---|---|---|
| ***Age*** | | | |
| *Mean* | *52.8* | *52.9* | *0.9898* |
| *≤40* | *6 (11.6)* | *4 (7.4)* | *0.7206* |
| *41-50* | *16 (30.8)* | *22 (40.8)* | *0.3419* |
| *51-60* | *18 (34.6)* | *19 (35.2)* | *0.7343* |
| *61-70* | *10 (19.2)* | *4 (7.4)* | *0.9585* |
| *>70* | *2 (3.9)* | *5 (9.3)* | *0.0718* |
| ***Ethnicity*** | | | |
| *White Scottish* | *26 (50.0)* | *21 (38.9)* | *0.3284* |
| *White English* | *6 (11.6)* | *8 (14.8)* | *0.7759* |
| *Other* | *1 (1.9)* | *0* | *0.4906* |
| *Not disclosed* | *19 (36.5)* | *25 (46.3)* | *0.3308* |
| ***Menopausal status*** | | | |
| *Premenopausal* | *14 (26.9)* | *15 (27.8)* | *>0.9999* |
| *Perimenopausal* | *6 (11.5)* | *7 (13.0)* | *>0.9999* |
| *Postmenopausal* | *30 (57.7)* | *30 (55.6)* | *0.8428* |
| *Unknown* | *2 (3.9)* | *2 (3.7)* | |
| ***Presenting complaint*** | | | |
| *PMB* | *28 (53.9)* | *26 (48.2)* | *0.5672* |
| *HMB* | *13 (25.0)* | *16 (29.6)* | *0.6657* |
| *IMB* | *6 (11.5)* | *8 (14.8)* | *0.7759* |
| *Subfertility* | *2 (3.9)* | *0* | *0.2383* |
| *Incidental finding* | *3 (5.8)* | *4 (7.4)* | *>0.9999* |
| ***Parity*** | | | |
| *Nulliparous* | *16 (30.8)* | *7 (13.0)* | ***0.0308** |
| *1-4* | *29 (55.8)* | *40 (74.1)* | ***0.0222** |
| *≥5* | *2 (3.9)* | *1 (1.9)* | *0.6170* |
| *Unknown* | *5 (9.6)* | *6 (11.1)* | |
| ***BMI*** | | | |
| *Mean* | *37.9* | *38.3* | *0.8661* |

| | | | |
|---|---|---|---|
| *21-25* | *5 (9.6)* | *2 (3.7)* | *0.4425* |
| *26-30* | *2 (3.8)* | *5 (9.3)* | *0.1964* |
| *31-35* | *8 (15.4)* | *2 (3.7)* | *0.1411* |
| *36-40* | *9 (17.3)* | *8 (14.8)* | *0.5687* |
| *>40* | *6 (11.5)* | *8 (14.8)* | *0.2149* |
| *Unknown* | *22 (42.3)* | *29 (53.7)* | |
| ***Co-morbid factors^*** | | | |
| *Diabetes mellitus* | *12* | *12* | *>0.9999* |
| *PCOS* | *7* | *1* | ****0.0264*** |
| *HRT use* | *4* | *7* | *0.5283* |
| *Tamoxifen use* | *4* | *4* | *>0.9999* |
| *≥ 2 of above* | *2* | *1* | *0.6060* |

^Complete co-morbid factor information unavailable for n=5 EIN and n=3 HwA patients. Statistical analysis performed using the 2-sided Fisher's exact test to determine statistical differences between the categorical data and a two-tailed unpaired t-test used to compare the means of the continuous data, for those with EIN and those with HwA. *p<0.05.

**Supplemental Table 4. Summary of ARID1A immunoreactivity and comparison to diagnosis using WHO and EIN criteria**

| ARID1A | Subtype null | EIN | HwA | P value |
|---|---|---|---|---|
| N number | | 51 | 54 | |
| Positive (%) | | 45 (88.2) | 54 (100) | 0.011* |
| Null (%) | | 6 (11.8) | 0 | |
| | Isolated glands | 1 (2.0) | 0 | 0.486 |
| | Confluent glands | 4 (7.8) | 0 | 0.052 |
| | All glands null | 1 (2.0) | 0 | 0.49 |

^Subgroups of Null ARID1A immunoreactivity included for comparison. HwA = Hyperplasia without Atypia, EIN = Endometrial Intraepithelial Neoplasia. Percentages in brackets. Statistical analysis performed using a Fisher's exact test, 2-sided. *p<0.05.

**Supplemental Table 5 Patient characteristics and numbers of endometrial hyperplasia patients broken down by cluster group following immunostaining.**

| | **Cluster 1** | **Cluster 2a** | **Cluster 2b** | **Cluster 3** |
|---|---|---|---|---|
| | N=12 | N=4 | N=54 | N=35 |
| **Age** *(average)* | *51.4* | *51.3* | *53.2* | *52.2* |
| <40 | 3 | 0 | 3 | 4 |
| 41-50 | 2 | 2 | 22 | 12 |
| 51-60 | 5 | 1 | 20 | 11 |
| 61-70 | 1 | 1 | 4 | 8 |
| >70 | 1 | 0 | 5 | 0 |
| **BMI** *(average)* | *35.33* | *34.5* | *38.7* | *39.1* |
| 18.5-25 | 1 | 0 | 2 | 2 |
| 26-30 | 0 | 0 | 5 | 0 |
| >30 | 5 | 2 | 18 | 16 |
| unknown | 6 | 2 | 29 | 17 |
| **Post-menopausal** | | | | |
| Yes | 5 | 2 | 31 | 21 |
| No | 7 | 2 | 23 | 14 |
| **Presentation** | | | | |
| PMB | 5 | 2 | 29 | 17 |
| HMB | 4 | 1 | 14 | 10 |

| | | | | |
|---|---|---|---|---|
| other | 3 | 1 | 12 | 8 |
| *Nulliparous* | *3* | *2* | *8* | *11* |
| *Parous* | *7* | *1* | *40* | *22* |
| *Unknown* | *2* | *1* | *6* | *2* |

Body max Index (BMI) categorised according to the World Health Organisation (WHO) categories. PMB = Postmenopausal bleeding, HMB = Heavy menstrual bleeding.
